# ALEX: Automatic Language Explanations for Interpreting Treatment Effects via Multi-Agents

**DOI:** 10.64898/2026.04.23.26351510

**Authors:** Mingyu Lu, Chanwoo Kim, Younghoon Kwon, Nathan J. White, Su-In Lee

## Abstract

Precision medicine requires understanding how general treatment effects from randomized clinical trials should be applied to individual patients. Machine learning methods have shown some promise for estimating patient-level treatment effects, however, their clinical utility remains limited because they often fail to explain why responses to a given treatment vary across individuals. Here we present ALEX, an explainable AI (XAI)-driven, multi-agent framework that addresses this interpretability gap by translating the variables that drive precision predictions into data-grounded, natural-language clinical explanations. ALEX first independently identifies important subgroup treatment effects present in randomized clinical trials and then hands them to large language model (LLM) agents to produce contextualized and scrutinized clinical insights. Across five landmark randomized controlled trials, ALEX outperformed existing agentic methods on treatment explanation quality metrics consistent with the biomedical literature that aligned with blinded reviews by specialist physicians across the United States and Taiwan. In empirical case studies, ALEX provided key interpretable insights, such as identifying baseline glucose level as explaining the divergent findings between the ACCORD-BP and SPRINT trials, and proposed age as a novel and key effect modifier for pre-hospital tranexamic acid efficacy after trauma. These findings suggest that ALEX can help translate treatment effect heterogeneity into clinically grounded explanations that enable precision medicine.

## Introduction

Understanding whether, for whom, and why a healthcare intervention or treatment works is a central challenge in clinical decision-making, where healthcare professionals are limited to using available evidence provided in broad terms towards personalized treatment choices for individual patients ^1^. Recent studies have applied causal machine learning to estimate individual patient treatment effects ^2–4^—that is, changes in treatment outcomes depending on patient characteristics, such as age and sex—rather than simply predicting outcomes as in standard machine learning. Although these approaches can estimate individual treatment effects, they do not explain the underlying rationale or offer clinical insight, which limits their utility in medicine, where clinician trust, transparent reasoning, and informed decision-making are critical.

Explainable AI (XAI) methods have therefore been adopted to improve the transparency of these “black-box” models ^5^. In particular, feature attribution methods, such as Shapley value ^6^, are increasingly used to quantify the influence of patient characteristics on a model’s prediction ^7^. Although these methods have shown promise in clinical settings, an important gap remains ^8 9^. These methods produce numerical attribution signals but do not, on their own, provide clinically meaningful reasoning. For clinical use, such signals must be translated into human-interpretable rationales that are grounded in biomedical knowledge and aligned with clinical guidelines.

Early efforts to translate these XAI outputs into natural language have largely produced superficial descriptions of feature–outcome correlations (for example, simply stating that an increase in a specific patient variable alters the predicted clinical outcome by a certain margin) ^10^. Such summaries fall short of the deeper insights needed to understand treatment-effect heterogeneity in clinical practice, where changes to a patient’s care plan must be supported by verified clinical reasoning. Addressing this gap requires computational methods that can synthesize biologically and clinically plausible rationales for why treatment responses vary across patient populations. Recent advances in multi-agent large language model (LLM) systems have shown promise for this type of synthesis ^11–13^. However, these general-purpose frameworks have not yet been established for clinical use. While agentic frameworks for medical decision-making exist ^14,15^, these agents have focused on diagnostic reasoning rather than on explaining treatment-effect heterogeneity of randomized clinical trials (RCT) which are widely considered to provide “gold standard” level of evidence regarding treatment effects ^16^. Taken together, these limitations highlight the lack of methods that can connect treatment-effect estimates to clinically interpretable, mechanism-informed explanations grounded in clinical data.

To overcome this interpretability barrier, we present **ALEX** (**A**utomatic **L**anguage **EX**planations for Interpreting Treatment Effects via Multi-Agents), an XAI-driven, multi-agent system for generating language explanations for treatment-effect heterogeneity. Rather than treating XAI output as a final endpoint, ALEX translates it into clinical rationale using natural language, enabling clinicians to understand and contextualize the model’s reasoning. In doing so, ALEX improves interpretability and transparency, thus improving trust. The framework comprises three agents: An XAI agent uses source data from a RCT via external tool calls to train an ensemble treatment effect estimator, compute Shapley values, and generate a summary report of important features influencing the overall treatment effect; an XAI-guided generative agent that converts these signatures into structured clinical explanations; and an independent critic agent that refines the generated explanations to reduce overstatements and hallucinations.

We then validated the generated natural-language explanations by designing a comprehensive, three-axis evaluation framework tailored to clinical safety standards. Specifically, to systematically penalize hallucinations and ensure validity, we assessed explanation quality through a strict sequential check based on clinical standards, automated PubMed evidence retrieval, and evaluation by clinical specialists. ALEX was benchmarked against existing methods using five landmark randomized controlled trials (RCTs), including CRASH-2^17^, IST-3^18^, ACCORD-Glycemia ^19^, ACCORD-BP ^20^, and SPRINT ^21^. Our results demonstrate that ALEX consistently outperforms existing methods in producing logically coherent, literature-corroborated, and clinically actionable rationales for identified subgroup treatment effects. Additionally, a blinded expert assessment by 19 specialist physicians across the United States and Taiwan further supported the clinical relevance and actionability of ALEX’s outputs.

Finally, to facilitate clinical transition, we demonstrate the framework’s ability to address two common hurdles present when applying RCT evidence to real-world practice: differences in patient characteristics and variation in clinical care settings. Specifically, we introduce cross-cohort attribution analysis in ALEX, which compares patients within one cohort to baseline patients from other cohorts. Using this approach, we identify features that help explain the conflicting results of the ACCORD-BP and SPRINT trials. In a trauma-care case study, using a local trauma patient registry as the baseline, ALEX identifies key features associated with the efficacy of pre-hospital tranexamic acid (TXA), recapitulating patterns consistent with an independent randomized trial. Together, these results suggest that ALEX can help bridge the gap between estimated treatment-effect heterogeneity and clinically grounded explanations of why interventions may work differently across populations and care settings.

**Fig. 1.**
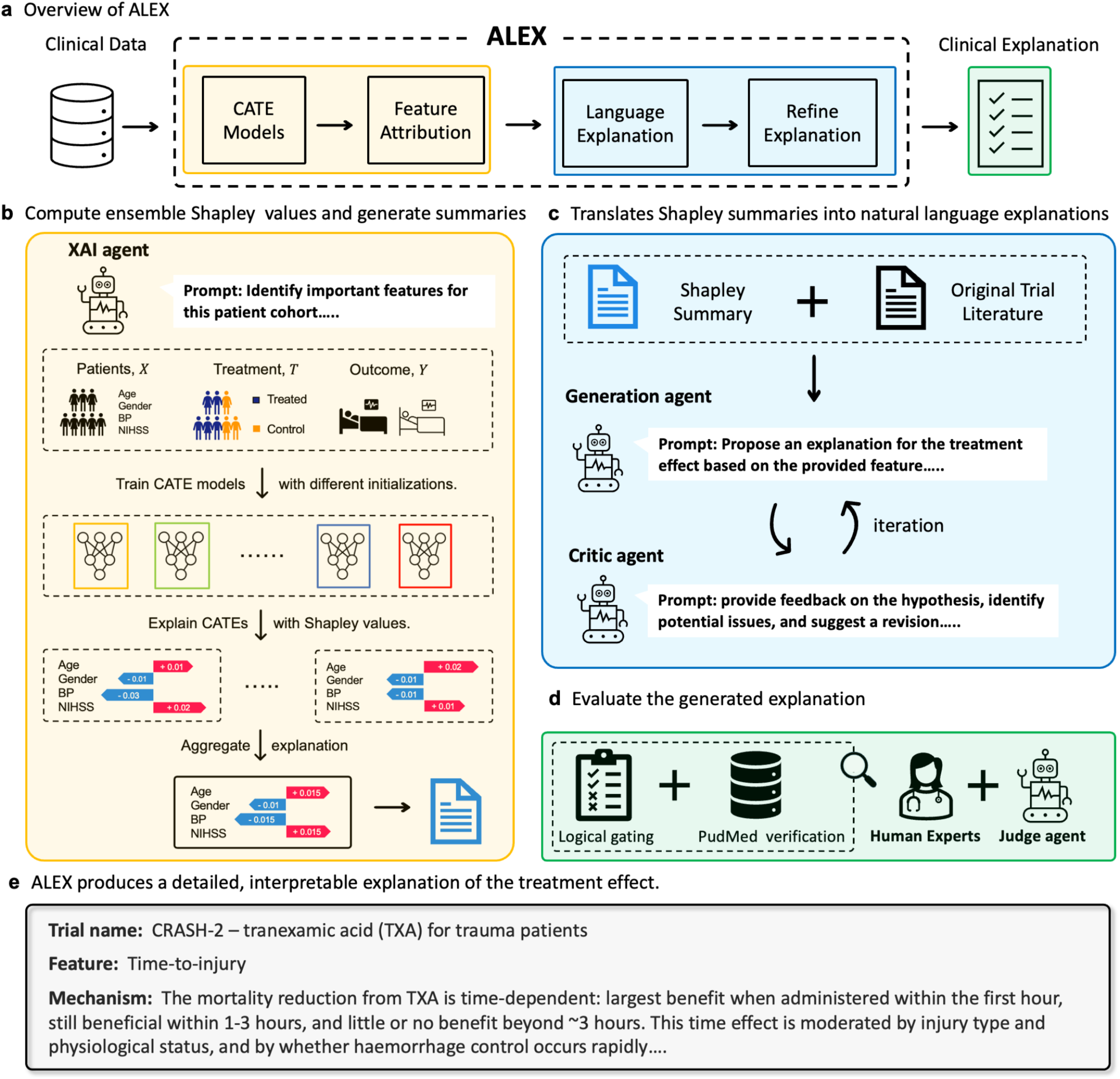
Framework of ALEX for clinical explanation generation for treatment effects. Overview of the ALEX framework. **a**, Overview of the ALEX framework, which translates clinical trial data into verified natural-language explanations. **b**, XAI agent trains an ensemble conditional average treatment effect (CATE) model and computes Shapley values. Patient covariates (X), treatment (T), and outcomes (Y) are used to train CATE models. Shapley values quantify the directional impact (red, positive; blue, negative) of individual features and are aggregated into a textual summary. **c**, Natural-language explanation generation and refinement. A generation agent uses the contextual summary together with the original trial literature to generate mechanistic explanations. A secondary verifier agent then critiques and iteratively refines these hypotheses. **d**, Evaluation via logic-based gating and PubMed validation, performed by an independent judge and clinical experts. **e**, Example explanation generated from ALEX.

## Results

In this section, we show that ALEX, a multi-agent framework that interprets treatment effects and generates corresponding language explanations, produces reliable outputs. ALEX comprises three core components: an XAI agent, a generation agent, and a critic agent. We first demonstrate that the XAI agent, which combines ensemble CATE estimation with Shapley values, provides rigorous, high-fidelity explanations at both the cohort and individual levels. Second, to assess the clinical utility and scientific validity of the resulting language explanations, we evaluated the generated explanation through a protocol involving an LLM-as-a-judge and human clinical experts. Overall, our findings indicate that ALEX consistently reduces hallucination and generates data-backed explanations.

### ALEX identifies effect modifiers across synthetic and clinical datasets with XAI agent

The first stage of ALEX identifies key patient features that influence individual treatment effects. We automated this process using an XAI agent to train an ensemble of conditional average treatment effect (CATE) models and compute Shapley values for feature attribution (Fig. 2a). We evaluated the quality of these attributions across two complementary axes: (i) how accurately attributions align with the model’s internal decision-making (i.e., explanation faithfulness), and (ii) how well the method recovers features known to influence treatment effect (i.e., effect modifier identification).

**Fig. 2.**
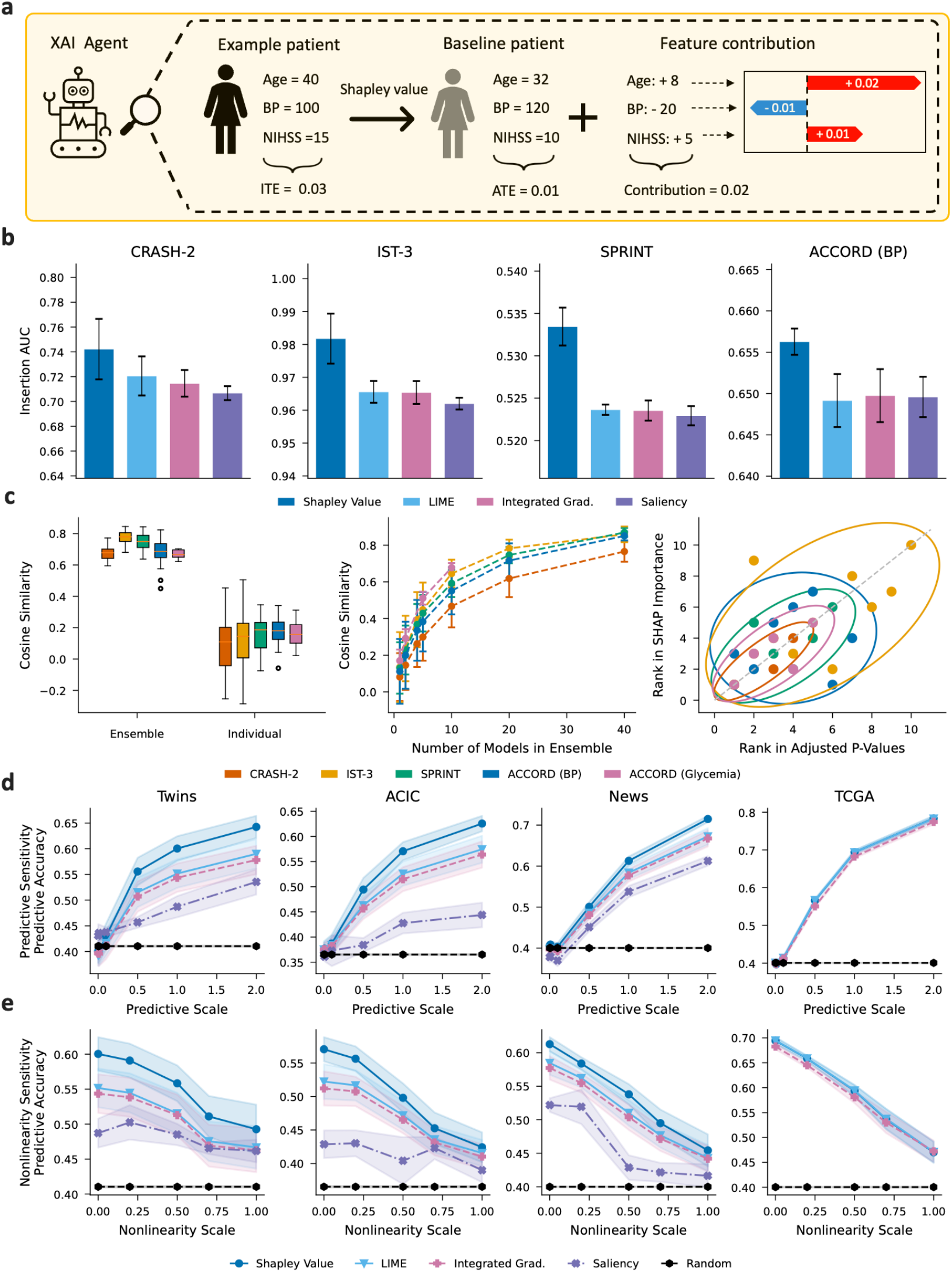
Ensemble Shapley value identifies effect modifiers in semi-synthetic and real-world datasets. **a**, XAI agent decomposes feature contributions for an example patient with Shapley value. **b**, Local fidelity: insertion AUC (pseudo-outcome, doubly-robust) across four datasets, CRASH-2, IST-3, SPRINT, and ACCORD (BP). Bars show mean AUC over seeds; error bars denote SEM. Higher values indicate that features identified as important by the method improve predictive performance when included first. **c**, Consistency of ensemble explanations. (Left) Cosine similarity of feature attributions for ensemble-averaged versus individual-model explanations. (Middle) Cosine similarity as a function of the number of models in the ensemble. (Right) Concordance between Shapley importance ranks and interaction *p*-value ranks across clinical datasets, with 95% confidence ellipses. **d, e**, Attribution accuracy of predictive features on semi-synthetic benchmarks, including Twins (*d*=8), ACIC (*d*=10), News (*d*=20), and TCGA (*d*=20) where *d* is the number of predictive features (effect modifiers). The *y*-axis reports predictive-feature attribution accuracy; the *x*-axis varies **(d)** predictive scale or **(e)** nonlinearity scale of the predictive features. Shaded bands show ±1 SEM across seeds.

To assess explanation faithfulness, we used standard insertion and deletion metrics, along with global distillation (Methods). Specifically, when evaluating the insertion metric across RCT datasets (Table 2), Shapley values yielded the highest normalized area under the curve (AUC) (Fig. 2b). Compared to alternative methods, including LIME and Integrated Gradients (IG), Shapley values isolated the specific features governing the underlying model’s behavior with higher fidelity. Furthermore, the ensembling strategy reduced the attribution variance typical of single-model explanations, increasing the mean cosine similarity from 0.15 for individual models to 0.80 for the ensemble (Fig. 3c-left, middle).

**Fig. 3.**
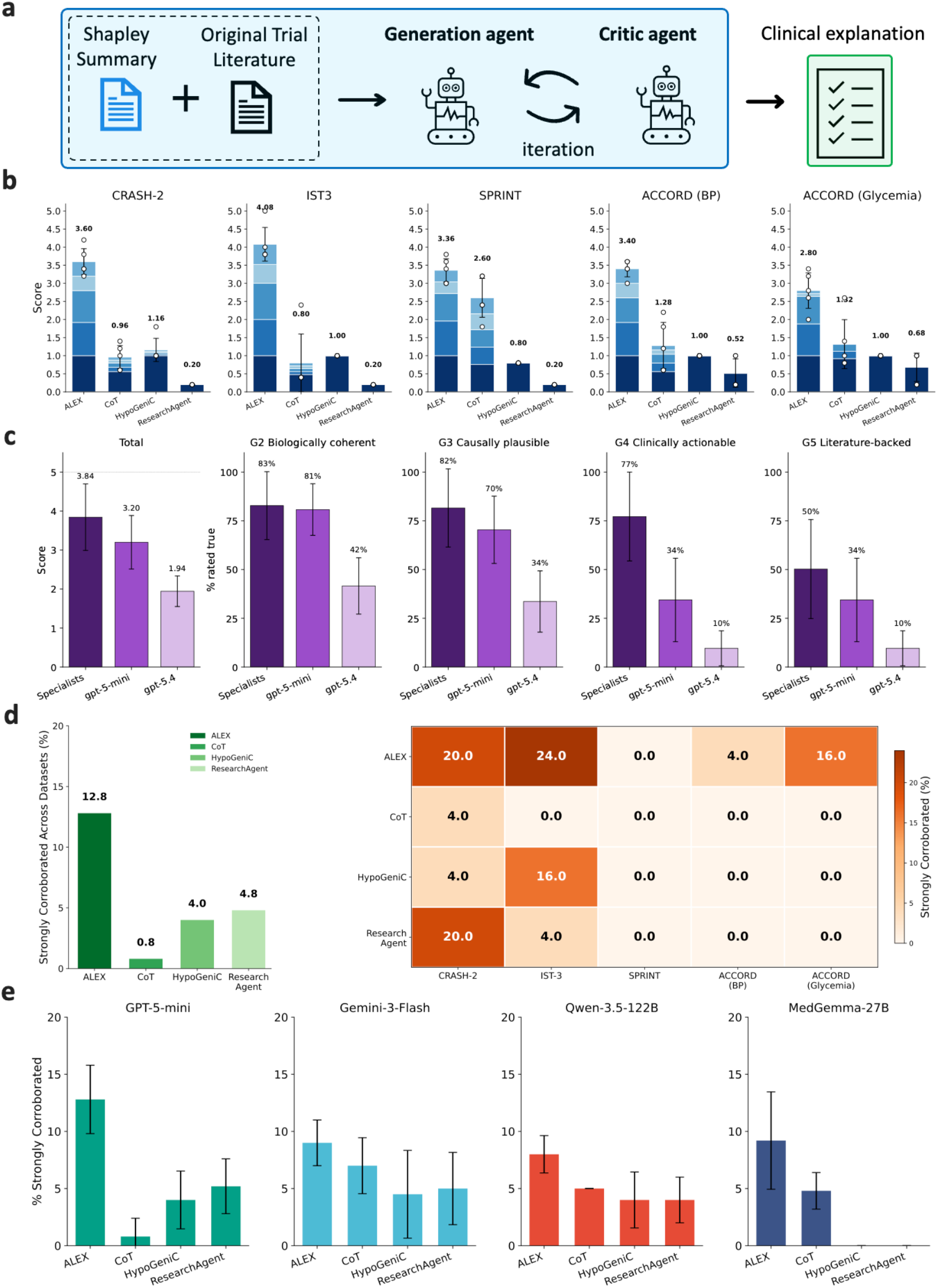
ALEX generates faithful and literature-corroborated clinical hypotheses across diverse datasets and base models. **a**, ALEX generates clinical explanations with XAI outputs and trial literature with generation and verifier agents. **b**, Mean logic-gate scores across datasets for each method evaluated by GPT-5-mini. Stacked bars represent performance across five criteria: data-grounded features (G1), logical coherence (G2), causal plausibility (G3), literature concordance (G4), and clinical actionability (G5). Dots indicate individual dataset scores, and error bars denote s.d. across datasets. **c**, Mean overall logic-gate scores and gate-specific scores across datasets for each method, as evaluated by specialist physicians, GPT-5-mini, and GPT-5.4. **d**, Strongly corroborated attribution rates across datasets. The left panel shows the mean percentage of hypotheses with robust PubMed support across all datasets for each method; the right panel details dataset-specific corroboration rates. **e**, Strongly corroborated attribution rates across various LLM base models, demonstrating ALEX’s architectural robustness.

For the second axis, we quantified the ability of these attribution methods to identify effect modifiers (i.e., predictive features) by measuring the average proportion of scores allocated to these variables. We initially evaluated four unconfounded semi-synthetic benchmarks (Section A), where the ground-truth effect modifiers are known. Across these datasets, in settings where the treatment effect correlates linearly with predictive features (scale = 1.0), Shapley values yielded accuracies up to 0.618 ± 0.011. Shapley values surpassed LIME (0.568 ± 0.013), Integrated Gradients (0.569 ± 0.013), and Saliency methods (0.484 ± 0.016) (Fig. 2d). In more challenging settings where the treatment effect is modeled as a non-linear function (scale = 0.5), Shapley values (0.430 ± 0.011) still maintained higher accuracy than all baselines (Fig. 2e).

Finally, we verified the XAI agent’s outputs against established real-world trial outcomes by evaluating its performance across the five RCT datasets. Specifically, we measured the concordance between Shapley value-based ranking of the features and their corresponding ranking based on reported subgroup interaction *p*-values from the original trials. Using Spearman’s rank correlation (*ρ*), we observed strong alignment across all trials with established effect modifiers (CRASH-2, IST-3, SPRINT, and ACCORD-Glycemia), with correlations ranging from *ρ* = 0.54 to *ρ* = 0.85 (Fig. 2c-right). Conversely, in trials lacking established effect modifiers and exhibiting inconclusive overall treatment effects, such as ACCORD-BP, the attributions appropriately reflected this absence, yielding a near-zero correlation (*ρ* = 0.05, *n* = 7).

### ALEX translates Shapley values into logical natural language explanations

While the XAI agent isolates the features driving the causal model’s predictions, these raw attribution scores do not provide actionable guidance. To bridge this gap, ALEX employs two agents: a generation agent first translates global Shapley summaries into natural-language narratives, which a subsequent critic agent evaluates and refines.

To ensure generated explanations meet rigorous clinical standards, we implemented a sequential logic-gating framework evaluated by an independent LLM judge. This step-by-step system requires explanations to pass each binary criterion before advancing. The resulting logic-gating score ranges from 0 to 5, where 0 indicates failure at the first gate, and 5 indicates that the explanation passed all five gates. The five gates are designed to ensure that outputs reflect coherent, data-grounded clinical reasoning rather than hallucinations (Methods). Using this framework, we compared the explanations (*n* = 25 per cohort per method) generated by ALEX and established baselines. Baselines include HypoGeniC ^13^, which derives explanations from patient-level outcome prediction, as well as literature-based approaches such as ResearchAgent^12^ and simple CoT prompting based on the original trial metadata.

Under this gating score evaluation (from 0 to 5), ALEX consistently outperformed existing methods across the clinical datasets, achieving an average total logic gate score of 3.61 ± 0.45, compared to CoT (1.41 ± 0.93), HypoGeniC (0.99±0.20), and ResearchAgent (1.00) (Fig. 3a). Table 1 presents representative qualitative explanations generated by each method. Subsequent gate-level analysis revealed distinct failure modes for the baseline methods. Despite having access to the original trial literature, the standard CoT and ResearchAgent approach yielded lower feature faithfulness (G1 = 0.59 ± 0.15), generating hypotheses that were coherent and plausible but based on patient characteristics, such as CT findings, not present in the original data; On the other hand, while HypoGeniC, a data-driven approach, avoided these feature hallucinations by using patient data directly, it did not pass downstream logic checks, yielding correlative statements (e.g., “higher X leads to a worse outcome”) rather than logically coherent descriptions (G2 = 0.01 ± 0.04). By integrating data-grounded XAI inputs with a generator-verifier agent to enforce clinical logic, ALEX successfully overcame both limitations. ALEX remained data-grounded (G1 = 1.00) while generating coherent (G2 = 0.95 ± 0.09) and causally plausible (G3 = 0.83 ± 0.17) interpretations of the observed treatment effects. Further examples of ALEX’s generated explanations are provided in Section G.

**Table 1.** Qualitative 5-gate evaluation of generated clinical explanation for tranexamic acid (TXA) efficacy in the CRASH-2 trial. All frameworks were tasked with interpreting the heterogeneous treatment effects of TXA on mortality. Generated hypotheses were evaluated across our sequential 5-gate framework: G1 (Feature Faithfulness), G2 (Logical Coherence), G3 (Causal Plausibility), G4 (Clinical Actionability), and G5 (Literature Concordance). The score is computed as the cumulative product across gates, summed over the sequence, so that failure at an earlier gate prevents credit from being assigned to later gates.

| Method | Proposed Clinical Explanation | Logic Gate Evaluation |  |  |  |  | Score |
| --- | --- | --- | --- | --- | --- | --- | --- |
|  |  | G1 | G2 | G3 | G4 | G5 |  |
| Chain-of-Thought (CoT) | TXA reduces bleeding-related mortality mainly in patients with objective **evidence of hyperfibrinolysis at presentation**, while patients with fibrinolytic shutdown derive little or no benefit (and may be harmed).<br>*Fails G1 (Feature Faithfulness): Presence/absence of active hyperfibrinolysis (physiologic phenotype measured by viscoelastic tests or biomarkers) was not measured in CRASH-2.* | × | ✓ | ✓ | × | × | 0 |
| HypoGeniC<sup>13</sup> | In trauma patients **aged <25 years**, tranexamic acid (TXA) reduces 28-day/in-hospital mortality compared with no TXA.<br>*Fails G2 (Logical Coherence): Grounded in data, but relies on shallow demographic correlations.* | ✓ | × | ✓ | × | × | 1 |
| ResearchAgent<sup>12</sup> | Patients with **different injury time levels** may show heterogeneous benefits due to variation in baseline disease burden. (variant 8)<br>*Fails G2 (Logical Coherence): Grounded in data, but the proposed explanation is vague and tautological.* | ✓ | × | × | × | × | 1 |
| ALEX (ours) | **Early post-injury period (injury time to treatment)** is characterized by heightened fibrinolytic activation (tissue plasminogen activator surge) and fragile nascent clots; giving TXA within the early window (especially 1 hour) prevents plasmin-mediated clot breakdown and reduces exsanguination risk.<br>*Passes all gates: the explanation relies on a feature present in the dataset and connects a specific clinical threshold to a plausible mechanism supported by prior literature.* | ✓ | ✓ | ✓ | ✓ | ✓ | 5 |

### Specialist evaluation of ALEX explanations via logic gating

To further assess the real-world clinical utility of ALEX, we conducted a blinded review with board-certified physicians (*n* = 19) across five specialties relevant to the RCTs evaluated. (Table S.12). These experts evaluated a subset of ALEX-generated explanations (five per cohort) using the same sequential logic-gating criteria. The physician ratings confirmed the clinical plausibility of ALEX’s outputs, yielding a mean overall score of 3.84 ± 0.33 (Fig. 3c, leftmost panel). As shown at the individual gate level (Fig. 3c, panels 2–4), physicians assigned high binary gating scores for logical coherence (G2: 0.83 ± 0.24), causal plausibility (G3: 0.82 ± 0.23), and clinical actionability (G4: 0.77 ± 0.33).

When comparing expert ratings with LLM-as-judge evaluations, we observed consistent rankings of scores. Both human experts and LLM judges ranked hypotheses from the CRASH-2 and IST-3 cohorts the highest (with mean physician scores of 4.7 for both) and those from the ACCORD-glycemia cohort the lowest. We also found that specialists assigned higher scores for clinical actionability (G4: 0.77), whereas LLMs generally assigned lower scores (0.34 for GPT-5-mini and 0.10 for GPT-5.4, respectively). Notably, overall physician ratings were systematically higher than the automated scores (e.g., from GPT-5-mini and GPT-5.4), indicating that LLM judges are more stringent evaluators than human experts. This effect was particularly pronounced in stronger models, such as GPT-5.4, which consistently assigned lower logic-gating scores than both GPT-5-mini and the human specialists. These findings suggest that an LLM-as-judge framework can function as a conservative gatekeeper for evaluating clinical explanations. More broadly, the blinded physician review provides independent validation that ALEX generates plausible and actionable explanations, reinforcing its potential utility in real-world clinical scenarios.

### ALEX generates evidence-backed insights using multi-agent system

To evaluate the clinical validity of the generated explanations, we assessed their concordance with peer-reviewed literature. To support evaluation at scale, we adapted recent work on LLM-assisted automated systematic review ^22,23^. For each explanation, an article retriever agent first reformulated the explanation using the PICO (Population, Intervention, Comparison, and Outcome) framework ^24^ and then translated the resulting PICO elements into a MeSH-based PubMed search query to retrieve up to 30 articles via the NCBI API. Retrieved abstracts were then reviewed by an independent evaluator agent, GPT-5-mini, and classified into one of six evidence tiers, reflecting the strength of the empirical support. We applied this evaluation to five explanations per method and dataset (Methods).

As shown in Fig. 3d (left panel), ALEX yielded the highest proportion of explanations with strong literature support, achieving a 12.8% corroboration rate across datasets. This surpassed HypoGeniC (4.0%), ResearchAgent (4.8%), and standard CoT (0.8%). Overall, data-driven approaches (ALEX and HypoGeniC) produced hypotheses with higher PubMed alignment than methods lacking access to patient-level data (CoT and ResearchAgent).

An analysis of the individual cohorts revealed that ALEX successfully recovered published subgroup–treatment interactions. These include the time-to-injury dependency for TXA in CRASH-2^25^, baseline HbA1c and blood pressure control in ACCORD-Glycemia ^26^, and an age-dependent effect of thrombolysis in the IST-3 cohort ^27^. In the SPRINT cohort, ALEX proposed an explanation consistent with the primary study’s initial findings; however, the evaluator agent retrieved subsequent analyses refuting this hypothesis ^28^, resulting in a lower corroboration rate (4%). A similarly low corroboration rate was observed for the ACCORD-BP trial, likely reflecting the study’s originally inconclusive primary treatment effect, even though ALEX identified a specific age-mediated explanation supported by later literature ^29^.

Additionally, to further assess the generalizability of the proposed framework, we evaluated ALEX and other baselines using a range of LLMs. ALEX consistently outperformed other frameworks in generating strongly corroborated explanations, regardless of the underlying backbone (Fig. 3e). Overall, proprietary models (e.g., GPT-5-mini ^30^ and Gemini-3-Flash ^31^) yielded higher corroboration rates than open-weight alternatives (e.g., Qwen-3.5 122B ^32^). Notably, the domain-specific MedGemma 27B ^33^, despite being fine-tuned on medical corpora, underperformed relative to these general-purpose models. HypoGeniC and ResearchAgent, in particular, degraded substantially when paired with MedGemma 27B. Although ALEX also showed some performance decline with smaller open-weight backbones, it remained the top-performing method overall.

Finally, to confirm the validity and stability of our automated evaluation pipeline, for PubMed corroboration, we assessed the agreement between the LLM and a physician on retrieved articles and their classifications. The exact agreement was 94.3%, with a Cohen’s kappa of 0.850 (Table S.14). For logic gating, we assessed the inter-agent reliability between the primary evaluator (GPT-5-mini) and a stronger model (GPT-5.4). Across all explanations generated by ALEX, the evaluators demonstrated high concordance. While exact categorical agreement across the six evidence tiers was 62%, adjusting for ordinal proximity, where models assigning adjacent tiers are appropriately credited, revealed substantial inter-judge reliability (quadratically weighted Cohen’s *κ* = 0.72; Gwet’s AC2 = 0.90).

### Applying ALEX to decipher treatment effects across RCTs when patients are different

A primary barrier to the generalizability of RCTs is the variation in patient characteristics that exist outside of the controlled RCT framework. We stress-tested ALEX’s capacity to identify these differences in the context of intensive blood pressure management, comparing two landmark trials with discordant results: SPRINT, which demonstrated reduced cardiovascular mortality in high-risk non-diabetic patients with intensive blood pressure management, and ACCORD-BP, which found no significant benefit of the same intervention in a cohort of patients with type 2 diabetes ^20,21^.

With its XAI agent, ALEX enables cross-cohort comparisons by simply adjusting the reference baseline for computing Shapley values. Specifically, we substituted the diabetic baseline of the ACCORD-BP cohort with representative non-diabetic individuals from the SPRINT cohort (Fig. 4a). This reframes the original question—*which features are important for ACCORD-BP individuals compared to the average ACCORD-BP population?* —to instead ask *which features are important for ACCORD-BP individuals compared to the average SPRINT population?* By adjusting the reference group accordingly, this approach supports tailored, context-specific interpretations across studies.

**Fig. 4.**
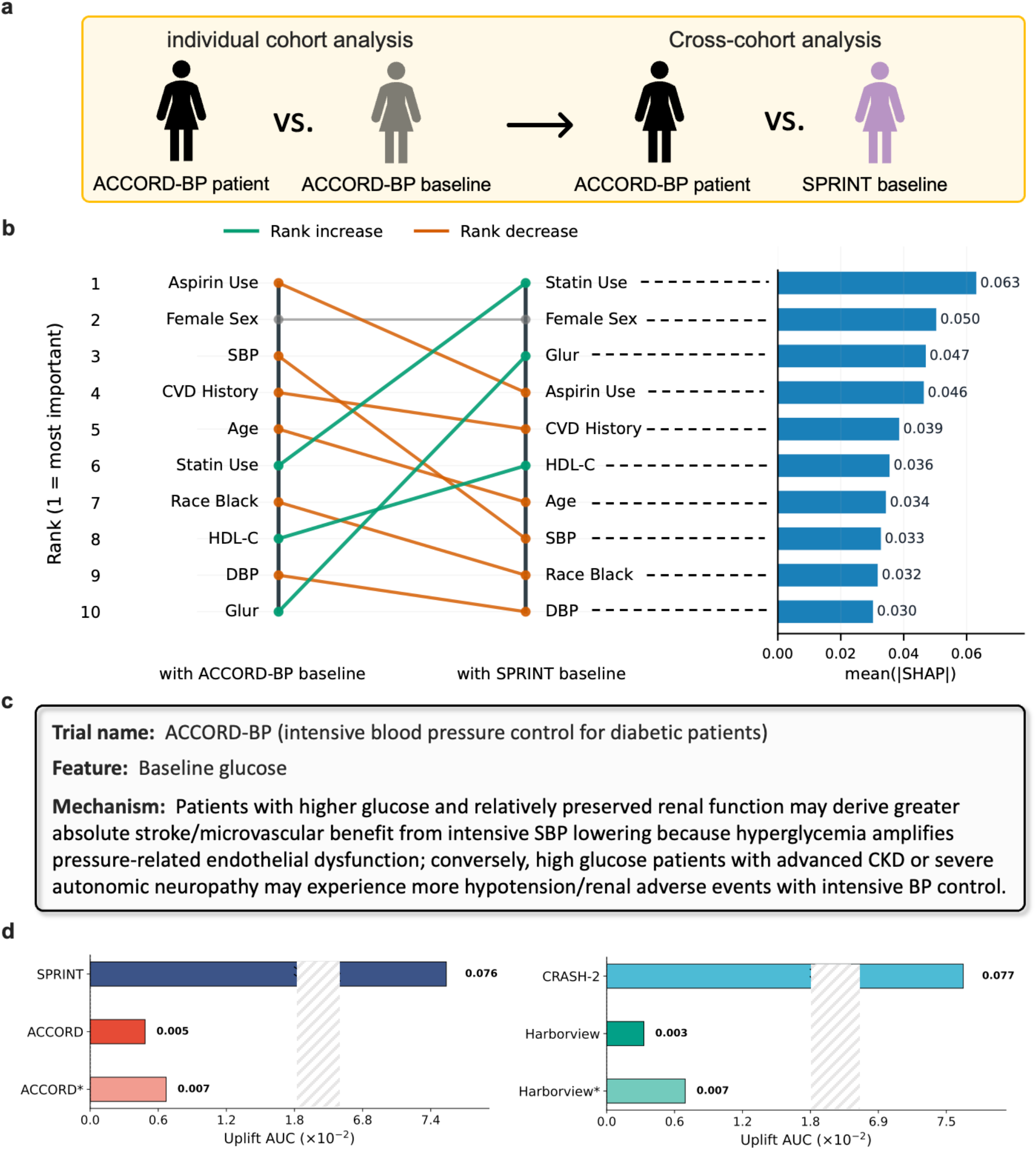
Cross-cohort analysis and treatment uplift evaluation. **a**, Overview comparing individual and cross-cohort analyses, achieved by swapping the average cohort baseline population during Shapley value computation. **b**, Comparison of top contributing feature ranks (left) and absolute average Shapley values (right) for ACCORD-BP patients. The analysis contrasts an individual cohort approach against a cross-cohort approach using SPRINT as the baseline. **c**, Representative explanation of the cross-cohort analysis utilizing the ALEX. **d**, Uplift scores (treatment gain) comparing the SPRINT and ACCORD-BP cohorts (left), and the CRASH-2 and Harborview trauma registry cohorts (right). * denote modified patient populations: the exclusion of individuals with glucose levels >160 mg/dL in ACCORD, and the exclusion of patients aged >65 years in the Harborview trauma registry.

Using this comparison, ALEX identified glucose-related features, particularly fasting plasma glucose (FPG), as important drivers of treatment-response heterogeneity between the trials (Fig. 4b). The hypothesis and verifier agents generated a candidate mechanistic explanation, suggesting that hyperglycemia-related endothelial dysfunction may contribute to this effect (Fig. 4c). An independent judge agent subsequently identified prior literature supporting glucose as a treatment-effect modifier ^34^, though the specific biological mechanism remains for further investigation. Importantly, glucose-related features did not emerge as prominent modifiers when the trials were analyzed independently (Fig. 4b, left panel), indicating that cross-cohort baseline adjustment was necessary to reveal this latent source of heterogeneity.

To further examine this signal, we analyzed the treatment benefit using uplift scores (Section B.3) among patients with varying glucose levels. As shown in Fig. 4 (d-left), the treatment effect efficacy, i.e., uplift score, for the original ACCORD-BP cohort was 5 × 10^−3^, substantially lower than that of the SPRINT cohort (7.6 × 10^−2^). However, among patients with FPG levels above 160 mg/dL—the median value in the ACCORD-BP cohort—and with preserved renal function, the estimated average treatment effect in ACCORD-BP increased by 40%. Together, these findings suggest that differences in glycaemic status may explain the divergent treatment effects observed across the two trials.

### Applying ALEX in an observational study to generate new explanations

Finally, we evaluated ALEX’s ability to generate actionable natural-language explanations in a more challenging scenario: when a proven treatment is applied to a different clinical setting. For this test, we considered the treatment of traumatic bleeding after injury using tranexamic acid (TXA), a drug that is used to stabilize blood clots to reduce bleeding after injury. Strong randomized data favor the use of TXA for trauma victims at risk of significant bleeding if given at hospital admission and within 3 hours of injury ^17^. Time from injury has emerged as having an important effect on TXA efficacy. Consequently, clinical practice has steadily crept towards using this drug faster, either at the scene of injury or during transport (pre-hospital), despite the lack of randomized evidence for its efficacy in this alternative practice setting. Using the original CRASH-2 data ^17^ alongside observational data from our local trauma center registry (Section A.2), we tasked ALEX with explaining the drivers of TXA efficacy in hospital (CRASH-2) versus pre-hospital (Harborview) settings.

In its generated explanation for the pre-hospital cohort (Section G.2), ALEX proposed that while established factors like time-to-injury and Glasgow Coma Scale (GCS) remain relevant, *age* emerges as a novel and critical effect modifier—a subgroup effect absent from the CRASH-2 in-hospital subgroup analysis. To empirically validate this discovery, we quantified the treatment benefit across age groups using uplift and Qini scores. For the pre-hospital cohort, the uplift score and qini score are 5 × 10^−4^ and −5 × 10^−4^, respectively. After excluding patients aged over 45 years in the pre-hospital settings, the scores increase to 5 × 10^−3^ and 8 × 10^−4^, respectively. Additionally, this age-dependent efficacy proposed by ALEX was recently observed and validated in the independent RCT PATCH trial of prehospital TXA for trauma ^35^. This result highlights the ability of ALEX to identify important treatment effects when randomized clinical trial data are applied to different clinical practice settings.

## Discussion

A fundamental challenge in precision medicine is identifying and understanding the underlying mechanisms of heterogeneous subgroup treatment effects found in RCTs. ALEX advances this goal by generating natural language explanations through a data-centric, agentic framework grounded in explainable AI. While existing literature-based agentic systems can sometimes hallucinate—generating plausible explanations that rely on clinical features absent from the underlying dataset—ALEX integrates a data-centric pipeline with LLM agents to effectively reduce such risk (Fig. 3a). By leveraging LLM agents to rigorously analyze, refine, and interpret these data- and model-grounded signals, ALEX moves beyond shallow correlative statements and provides clinically plausible explanations from model outputs that are more interpretable in clinical contexts (Table 1).

A core advantage of ALEX is its integration of computational tools into the XAI agent to identify important patient characteristics for further analysis. Unlike existing data-driven systems that rely on unvalidated zero-shot LLM reasoning for treatment effect estimation, the XAI agent grounds its outputs in a two-step procedure. First, it estimates treatment effects using an ensemble of advanced causal models ^2,36^. Second, it derives feature attributions using Shapley values ^3,9^. This process prioritizes the most influential patient characteristics and grounds the resulting explanations in patterns learned by the causal models. As shown in our evaluation (Fig. 2), the features prioritized by the XAI agent broadly aligned with established treatment-effect modifiers.

Building on this foundation, ALEX is uniquely positioned to explore and explain possible sources of discordance across major clinical trials. We demonstrated this capability by investigating the divergent outcomes of SPRINT^21^ and ACCORD-BP ^20^ regarding intensive blood pressure management (Fig. 4). ALEX independently identified baseline blood glucose as a key factor, thus our cross-cohort analysis provides a data-driven rationale for why the diabetic cohort in ACCORD-BP did not derive the same cardiovascular benefits observed in SPRINT and suggests a FPG threshold whereupon diabetic patients may derive benefit. Although retrospective and subject to cross-trial confounding, this insight demonstrates how ALEX can assist precision patient selection for intensive antihypertensive therapy and motivate targeted, prospective studies in populations with impaired glucose regulation. Crucially, ALEX moves beyond mere statistical subgroup identification; it generates structured, mechanistic hypotheses—such as the interplay between glucose-related physiology and renal hemodynamics—that may help prioritize subgroups and candidate mechanisms for prospective testing in future trials.

Despite these strengths, ALEX is constrained by the inherent limitations of its underlying tools, such as causal models, feature attribution methods, or generative agents. First, while these causal estimators perform well in the controlled setting of randomized trials, their reliability diminishes in observational studies where unobserved confounding can violate key identification assumptions ^3,4,37^, propagating bias directly into the generated clinical explanations. Second, although Shapley values isolate candidate modifiers, their ability to recover predictive features can weaken in the presence of highly complex nonlinear relationships ^38^. Third, the clinical fidelity of any multi-agent system is intrinsically bound by its foundation model. Consistent with other agentic architectures, we observed that explanatory robustness degrades with model size, increasing the risk of hallucinations even when a verifier agent is present. Finally, while our automated logic-gating procedure provides a scalable mechanism for comparing model outputs, it remains a proxy for true clinical utility. We observed that the LLM judges adhere rigidly to evaluation prompts, such as strictly crediting only explicitly stated mechanisms. In contrast, human clinicians naturally leverage their domain expertise to infer missing mechanistic links. Consequently, before any real-world deployment, the explanations generated by ALEX will require prospective, blinded validation by multidisciplinary expert panels to mitigate the risk of automation bias.

Looking forward, several avenues are critical for translating ALEX into routine clinical research and practice. Methodologically, extending this framework beyond controlled trial environments to observational electronic health record (EHR) data will require robust methods for mitigating unmeasured confounding, such as integrating instrumental variable-based attributions ^39^ and formalizing causal domain knowledge ^40^. Clinically, while our retrospective application to SPRINT and ACCORD-BP highlights its utility for hypothesis generation, prospective validation is necessary. Future studies should deploy ALEX within adaptive clinical trial designs to prospectively identify treatment-effect modifiers and enrich patient subgroups in real time. Finally, as multi-agent LLM systems evolve, research must prioritize “human-in-the-loop” evaluation paradigms. Designing interfaces that allow human experts to safely audit and refine AI-generated mechanistic hypotheses will be essential for mitigating automation bias and ensuring that these systems rigorously augment clinical domain expertise.

In conclusion, ALEX provides an XAI-driven, multi-agent framework for interpreting heterogeneous treatment effects. By integrating external tool usages with multi-agent LLMs, the approach converts quantitative estimates of treatment-effect heterogeneity into clinically contextualized, testable hypotheses. Although prospective validation and expert adjudication remain essential before any real-world use, ALEX may be valuable for trial interpretation, subgroup hypothesis generation, and the prioritization of candidate mechanisms for further investigation. As precision medicine advances, frameworks that improve the transparency and auditability of heterogeneous treatment-effect analyses may help strengthen the translation of trial data into clinically relevant research questions and precision therapies.

## Methods

### Datasets

We analyzed five publicly available randomized controlled trials (RCTs) with binary outcomes: IST-3 (intravenous alteplase), CRASH-2 (tranexamic acid), SPRINT (intensive blood-pressure control), ACCORD-BP (intensive blood-pressure control in type 2 diabetes), and ACCORD-Glycemia (intensive glycemia control). For each trial, we removed covariates with substantial missingness and one-hot encoded categorical variables. Key cohort characteristics, covariates, and endpoints are summarized in Table 2.

**Table 2.**
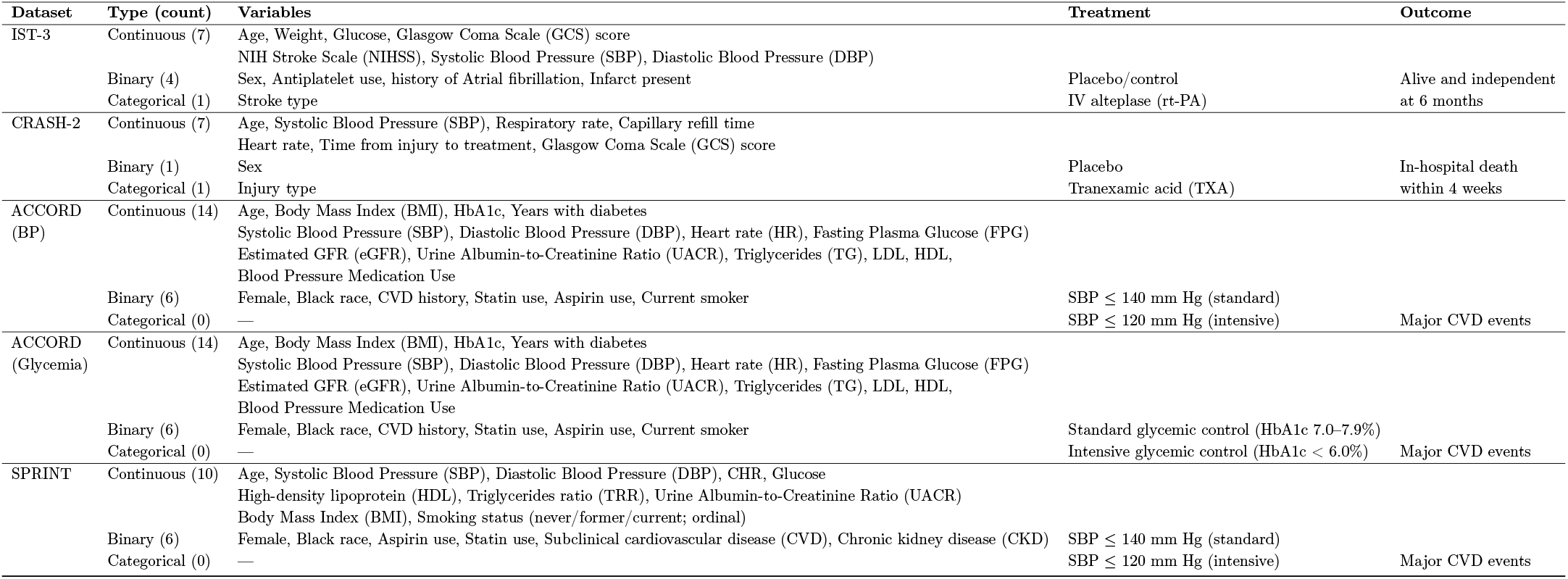
Curated dataset descriptions, including features, treatment, and outcome definitions.

#### IST-3

This trial evaluated intravenous alteplase (rt-PA) versus control in patients with acute ischemic stroke (*n* = 3,035)^18^. The dataset includes 7 continuous, 4 binary, and 1 categorical covariate (stroke type), with 6-month survival as the binary outcome.

#### CRASH-2

This trial tested tranexamic acid (TXA) against placebo in bleeding trauma patients (*n* = 20,211)^17^. The dataset includes 7 continuous, 1 binary, and 1 categorical variable (injury type), with in-hospital survival defined as the binary outcome.

#### SPRINT

SPRINT ^21^ (*n* = 9,361) compared intensive blood pressure control with a standard target. The dataset contains 12 continuous and 7 binary covariates, with the primary binary outcome defined as freedom from composite cardiovascular events.

#### ACCORD-Glycemia

The ACCORD trial ^19^ evaluated the effects of intensive versus standard glucose-lowering therapy in patients with type 2 diabetes (*n* = 10,251). The dataset includes 18 continuous and 7 binary covariates, evaluated against a protocol-defined primary cardiovascular composite outcome.

#### ACCORD-BP

ACCORD ^20^ evaluated intensive versus standard systolic blood pressure targets in patients with type 2 diabetes (*n* = 4,733). The dataset includes 18 continuous and 7 binary covariates, with a protocol-defined primary cardiovascular composite outcome.

In particular, SPRINT and ACCORD-BP enrolled overlapping patient populations but reported contrasting outcomes for intensive blood pressure control, offering a natural case study for whether interpretability methods capture clinically meaningful differences.

To complement these RCTs, we incorporated a retrospective cohort to assess performance in a real-world clinical setting. This retrospective cohort comes from the Harborview pre-hospital TXA registry (2007-2020) Section A.2, serving as a reference population for CRASH-2. We applied propensity-score matching ^41^ to construct a balanced sample of 240 patients (120 TXA, 120 controls) from trauma admissions. The dataset contains 6 continuous and 1 categorical variable, with survival as the outcome. Detailed variable definitions of the above datasets are provided in Table 2.

### Overview of ALEX

ALEX is an end-to-end framework that converts raw clinical trial data into testable language explanations. Conditioned on trial context (treatment, outcome, and population) and patient-level covariates, ALEX executes a three-step workflow. First, its XAI agent involves training an ensemble of conditional average treatment effect (CATE) estimators to model treatment-effect heterogeneity. Second, it summarizes the feature-level drivers of this heterogeneity using Shapley value computed across the ensemble. Finally, it employs generation and verification agents to translate these statistical attributions into structured, clinically actionable subgroup hypotheses.

#### XAI Agent for Attribution of Heterogeneous Treatment Effects

To identify clinically relevant effect modifiers, the XAI agent was designed to orchestrate external causal estimators and attribution tools rather than rely on free-form LLM reasoning alone. Under the potential–outcomes framework ^37^, the CATE for a binary treatment *T* ∈ {0, 1} and covariates *X* is

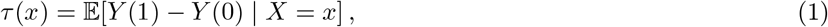

where *Y* (1) and *Y* (0) denote individual *i*’s potential outcomes under treatment and control. Because only one potential outcome is observed for each individual, *τ* (*x*) must be inferred from data. In the first stage, CATE was estimated using neural-network-based meta-learners ^1,38^, implemented with the `CATENets` package^1^. These methods estimate *τ* (·) by regressing learner-specific pseudo-outcomes:

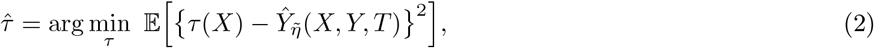

where 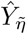 is constructed from nuisance functions 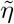. We use the DR-Learner ^42^ due to its robustness in identifying effect modifiers—features that are highly predictive of treatment benefit ^38,43,44^. In the second stage, feature attribution was obtained using Shapley values ^6^, computed through the `Captum` package^2^. Shapley values were chosen because they satisfy axiomatic properties, are additive, and can be aggregated naturally from individual-level attributions to cohort-level summaries. In practice, Shapley values were approximated by sampling with empirical feature means as baselines ^45^.

To improve robustness, we used attributions from an ensemble of CATEs because post hoc explanations from a single CATE model may be unstable owing to stochastic training dynamics, including random weight initialization and data batching. Let *τ*_*i*_ denote the *i*-th trained CATE model and 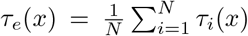 denote the ensemble predictor. For attribution methods satisfying linearity, ensemble attributions can be computed by averaging model-specific attributions:

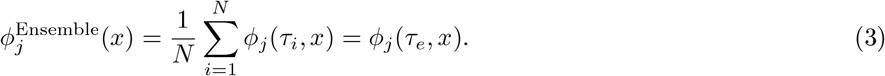

Implementation details for CATE, including model training, hyperparameter tuning, and Shapley value computation, such as baseline selection, are provided in Section B and Section C.

#### Translation of Attributions into Coherent Language Explanation

While Shapley values provide a rigorous quantification of feature contributions, they remain abstract numerical values that do not inherently convey human-understandable clinical concepts. To bridge this gap, ALEX utilizes a subsequent dual-agent LLM workflow to translate ensemble attributions into structured, schema-constrained hypotheses. We first condense the ensemble Shapley values, *ϕ*^Ensemble^, into a contextual summary, *S*, which integrates trial metadata (treatment, outcome, and population) with global importance ranks and effect directions. This summary serves as the input for a multi-stage generation and refinement process, where all outputs are strictly enforced by a predefined schema, *T*, to ensure robust evaluation and reproducibility.

#### Explanation Generation Agent

The generation agent functions as a clinical scientist, *G* : *S* → ℋ_raw_, prompted to propose multiple distinct mechanisms explaining each feature’s role as an effect modifier. To prevent superficial claims, the system prompt explicitly instructs the model to formulate precise treatment-response interactions. Specifically, the agent must propose a potential underlying mechanism, such as a specific biological or physiological pathway, that drives non-monotone effects and articulate which subgroups derive greater absolute benefit.

#### Critic Agent

A secondary critic agent, *V* : (ℋ_raw_, *S*) → ℋ_refined_, acts as an independent critic to optimize the initial hypotheses. Provided with the trial literature, the agent is prompted to identify mechanistic implausibilities, clarify clinical interpretations, and propose concrete validation steps by reformulating the raw hypotheses, ℋ_raw_. To preserve the structural diversity of the generation phase, the agent is programmatically constrained to revise the existing set without deleting any candidate mechanisms. This critique–revision cycle iterates for a prespecified number of rounds to yield the final hypothesis. Specific prompts for the XAI, generation, and verifier agents are provided in Section H.

#### Validation of CATE Models

Because the true CATE, *τ* ^∗^(*x*), is fundamentally unobservable for any given patient, we evaluate model performance using *pseudo-outcome surrogates* ^46,47^ (Section B.3) to approximate prediction error. Additionally, we quantify the clinical utility of the model’s treatment effect estimates using Qini and uplift curves ^48^ (Section B.3), which measure cumulative treatment effect gains across the sorted population. Model selection was performed via 5-fold cross-validation. The best-performing learner was subsequently designated as the target model for all downstream interpretability analyses. Beyond the DR-Learner, we benchmarked a diverse family of CATE estimators and inherently interpretable models (Section B.1).

#### Examining Interpretability Methods

To evaluate the quality of the generated explanations, we compared Shapley values against standard local attribution methods (Integrated Gradients ^49^, Saliency ^50^, SmoothGrad ^51^, LIME ^52^) and global importance baselines (LOCO ^53^, PermuCATE^54^) for CATE models (Section C.2). Explanations were evaluated along two critical axes:

##### Faithfulness to the Fitted Model

We assessed how accurately the explanations represent the underlying mechanics of the fitted CATE model, 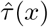. Local faithfulness ^55^ was evaluated via insertion and deletion metrics on 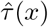 (Section C.3). Global sufficiency ^56^ was measured using top-*k* distillation, which quantifies the extent to which restricting the CATE model to only the highest-ranked features preserves the original predictions of 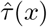 (Section C.3)

##### Identification of Predictive Features

A clinically useful explanation must successfully isolate true effect modifiers (predictive features) from purely prognostic (outcome-predictive) and irrelevant covariates. Because ground-truth contributions to treatment heterogeneity are unobserved in empirical data, we employed a dual evaluation strategy. First, we measured feature identification precision in semi-synthetic datasets where true effect modifiers are known by construction ^38^ (Section C.3). Second, for real-world trial datasets, we assessed concordance with external clinical evidence by calculating the association between our global feature rankings and the statistical significance of treatment–covariate interactions (interaction *p*-values) reported in the original trial literature.

#### Evaluation Metrics for Generated Hypotheses

We evaluated the generated feature-based treatment effect hypotheses along three complementary axes: (i) external evidentiary support, quantified via automated PubMed literature retrieval and stance classification; (ii) logical coherence and intrinsic validity, assessed by an independent LLM judge to penalize factual hallucinations; and (iii) clinical face validity, evaluated by blinded domain experts.

##### Sequential Logical Gating for Explanations

Because LLM hallucinations pose a critical risk in high-stakes clinical domains, we implemented a rigorous gating mechanism to evaluate hypothesis integrity. To evaluate the quality of the proposed hypothesis, we subjected each generated clinical hypothesis to a cascaded sequence of binary logic checks, *G*_*i*_ ∈ {0, 1}. The five sequential criteria are defined as follows:

- *G*_1_: Does the proposed feature actually exist in the dataset?
- *G*_2_: Is the proposed mechanism logically coherent? (e.g., penalizing vacuous explanations such as *higher values of X lead to more absolute benefit* ).
- *G*_3_: Is the mechanism causally plausible given the trial design? (e.g., penalizing the reliance on post-randomization or post-treatment variables).
- *G*_4_: Is the explanation clinically actionable?
- *G*_5_: Is the explanation supported by the retrieved literature?

While *G*_1_ and *G*_5_ are validated via deterministic programmatic and retrieval pipelines, respectively, we employed an independent LLM for *G*_2_, *G*_3_ and *G*_4_ with a set of positive and negative samples. Because downstream criteria depend on the fulfillment of upstream logic check, the final score *S*(*h*) rewards hypotheses that survive the sequence:

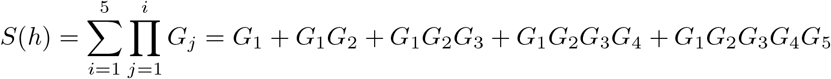

This multiplicative gating ensures that a hypothesis failing a fundamental check (e.g., *G*_1_ = 0 if the proposed feature does not exist in the dataset) receives no credit for any subsequent downstream criteria. The judge prompt for logic gating evaluation is provided in Section H. We utilized GPT-5-mini ^30^ as the primary backbone model for the judge, alongside additional experiments evaluating GPT-5.4 and Gemini-3-Flash ^31^. We evaluated agreement using weighted Cohen’s *κ*^57^ and Gwet’s AC2^58^.

##### Literature Discovery and Evidence Grounding

To quantify the evidentiary basis of each generated explanation, we developed a systematic retrieval pipeline utilizing the NCBI E-utilities API (https://www.ncbi.nlm.nih.gov/) and evaluated findings through a three-step approach:

- **Step 1: Systematic Retrieval**. An article retriever agent first reformulated each explanation according to the PICO framework and then decomposed it into a structured MeSH-based PubMed query restricted to clinical trials, reviews, and meta-analyses. We retrieve a representative sample of up to *k* abstracts (default *k* = 30) per explanation.
- **Step 2: Abstract Classification**. An independent LLM-as-judge evaluates each abstract’s relationship to the proposed explanation, assigning it to one of six stance categories: *support, weak support, prognostic, no interaction, conflicting*, or *irrelevant*.
- **Step 3: Explanation Classification**. Using the *k* labels and a decision-tree-based rule, we infer an overall evidence strength category: Strongly Corroborated, Weakly Corroborated/Debated, Likely No Interaction, Prognostic Main Effect Only, Refuted, or No Supporting Evidence.

The complete retrieve, judge prompt, and detailed evaluation criteria are provided in Section H. To ensure the accuracy of the LLM’s classifications, we also employed two physicians as human annotators and compared the LLM’s classifications with theirs using Cohen’s *κ*^59^ for generated explanations across methods.

##### Specialist Physician Evaluation

Finally, the ALEX-generated explanations were evaluated by a panel of 19 specialist physicians from North America (n=13) and Taiwan (n=6), with expertise in emergency medicine, neurology, internal medicine, endocrinology and metabolism, and cardiology. Each expert reviewed five sampled explanations per cohort, selected to align with their clinical specialty, along with the corresponding original cohort literature. The evaluating physicians were blinded to the source of the explanations and were not informed that the text was generated by ALEX. To ensure consistency, physicians received the identical questions and descriptions applied in the LLM-as-a-judge logic gating above. Specifically, they were asked to evaluate logical coherence (G2), causal plausibility and clinical actionability (G3), and literature backing (G4).

## Data Availability

UW Harborview data and physician survey data are not publicly available. All other data generated in this study are available online at the links below.

https://biolincc.nhlbi.nih.gov/home/

https://datashare.ed.ac.uk/handle/10283/1931

https://github.com/AliciaCurth/CATENets

## Data availability

The generation process for synthetic datasets is available on GitHub at https://github.com/AliciaCurth/CATENets. The IST-3 dataset is publicly accessible at https://datashare.ed.ac.uk/handle/10283/1931. The CRASH-2 dataset can be accessed at https://freebird.lshtm.ac.uk/index.php/available-trials/, with treatment allocations available upon request. Both the ACCORD and SPRINT datasets are available upon request at https://biolincc.nhlbi.nih.gov/home/.

## Code availability

The code for training, inference, and evaluation of the CATE models, XAI methods, ALEX, and other baselines used in this study is available at https://github.com/suinleelab/ALEX. The code is distributed under the BSD 3-Clause License. The model weights are provided and intended for non-commercial use only.

## Acknowledgement

We extend our gratitude to the Trial Collaborators at the London School of Hygiene & Tropical Medicine Clinical Trials Unit (University of London) for sharing CRASH-2 Trial data, and to the researchers in the Lee lab for their valuable discussions. We also thank the participating physicians and collaborating clinical institutions for their participation in the physician survey.

## Supplementary Information

## A Datasets

### A.1 Real-World RCTs

**IST-3 (The Third International Stroke Trial)**^**18**^ was a large, multicentre, randomized controlled trial designed to determine whether a broader range of stroke patients could benefit from intravenous recombinant tissue plasminogen activator (rt-PA) administered within six hours of acute ischemic stroke onset. The trial enrolled 3,035 patients across 156 hospitals in 12 countries, who were randomly assigned to receive either rt-PA or standard non-thrombolytic care. Functional outcomes were assessed at six months using the Oxford Handicap Scale (modified Rankin Scale). The results demonstrated that, in selected populations, the benefits of early thrombolytic therapy outweighed the associated risks.

The final curated dataset comprised 7 continuous variables (age, `weight`, `glucose`, `gcs-score-rand`, `nihss`, `sbprand`, `dbprand`), 4 binary variables (`gender`, `antiplat-rand`, `atrialfib-rand`, `infarct`), and 1 categorical variable (`stroketype`). The binary outcome variable represents six-month survival (`aliveind6`). For detailed information on inclusion criteria, participant demographics, and variable definitions, please refer to the original study ^18^ and its publicly available dataset at IST-3@DataShare.

**CRASH-2 (Clinical Randomisation of an Antifibrinolytic in Significant Haemorrhage)** ^**17**^ had the objective to evaluate the effect of early administration of tranexamic acid (TXA) on mortality, surgical intervention, and transfusion requirements in trauma patients with, or at risk of, significant bleeding. The randomized trial encompassed over 20,000 trauma patients. The intervention was TXA versus placebo. The major outcomes of interest were death in hospital within 4 weeks post-injury, need for blood transfusion, and surgical interventions.

The major discovery was that early administration of TXA reduced all-cause mortality without increasing the risk of vascular occlusive events. The curated analytic dataset comprised 9 covariates in total: 7 continuous (`iage`, `isbp`, `irr`, `icc`, `ihr`, `ninjurytime`, `igcs`), 1 binary (`isex`), and 1 categorical (`iinjurytype`, one-hot encoded for analysis). The binary outcome variable represented survival at hospital discharge. For detailed information on inclusion criteria, participant demographics, variable definitions, feature distributions, and trial methodology, please refer to its original study^17^ and the publicly available dataset at freeBIRD.

**SPRINT (Systolic Blood Pressure Intervention Trial)** ^21^ was a large, multicenter randomized controlled trial designed to assess the effects of intensive versus standard blood pressure control on cardiovascular outcomes and overall mortality in adults at elevated cardiovascular risk. The trial enrolled 9,361 participants with a systolic blood pressure of 130 mmHg or higher and at least one additional cardiovascular risk factor. Participants were randomly assigned to an intensive treatment group targeting a systolic blood pressure below 120 mmHg or a standard treatment group targeting below 140 mmHg. The primary composite outcome comprised myocardial infarction, non–myocardial infarction acute coronary syndrome, stroke, heart failure, or death from cardiovascular causes. The study demonstrated that intensive blood pressure control significantly reduced the rates of the primary composite outcome and all-cause mortality compared with standard treatment.

The dataset comprises 19 baseline variables: 12 continuous, 7 binary, and no categorical variables. The final analytic dataset includes 12 continuous variables (`age`, `sbp`, `dbp`, `n-agents`, `egfr`, `screat`, `chr`, `glur`, `hdl`, `trr`, `umalcr`, and `bmi`) and 7 binary variables (`female`, `race-black`, `smoke-3cat`, `aspirin`, `statin`, `sub-cvd`, and `sub-ckd`). The treatment variable (`intensive`) indicates the randomized intervention arm, and the outcome variable (`event-primary`) represents the occurrence of the composite cardiovascular endpoint. For detailed information on inclusion criteria, participant demographics, variable definitions, feature distributions, and trial methodology, please refer to its original study ^21^.

**ACCORD (Action to Control Cardiovascular Risk in Diabetes)** ^19,20^ was a large, multicenter randomized controlled trial designed to evaluate whether intensive control of cardiovascular risk factors reduces major cardiovascular events in adults with type 2 diabetes at high cardiovascular risk. The trial enrolled 10,251 participants and included randomized interventions for glycemic control, blood pressure control, and lipid management. The primary composite outcome was the first occurrence of nonfatal myocardial infarction, nonfatal stroke, or death from cardiovascular causes.

In the blood-pressure intervention, participants were assigned to intensive versus standard systolic blood-pressure targets, below 120 mmHg and below 140 mmHg, respectively. Intensive blood-pressure control did not significantly reduce the rate of the primary composite cardiovascular outcome compared with standard control ^20^. In the glycemic-control intervention, participants were assigned to intensive versus standard glucose-lowering strategies, targeting glycated hemoglobin levels below 6.0% versus 7.0–7.9%, respectively. Intensive glycemic control reduced some nonfatal cardiovascular events but was associated with increased all-cause mortality, leading to early termination of the intensive glycemia arm ^19^.

We curated two ACCORD-derived datasets. The ACCORD-BP dataset uses intensive versus standard blood-pressure control as the treatment variable, with the primary composite cardiovascular event as the outcome. The ACCORD-Glycemia dataset uses intensive versus standard glycemic control as the treatment variable, with the same primary composite cardiovascular outcome. Both datasets include baseline demographic, biochemical, clinical-history, and treatment-related covariates used to assess heterogeneous treatment effects.

The curated datasets from ACCORD comprise 18 continuous variables (`baseline-age`, `bmi`, `sbp`, `dbp`, `hr`, `fpg`, `alt`, `cpk`, `potassium`, `screat`, `gfr`, `uacr`, `chol`, `trig`, `vldl`, `ldl`, `hdl`, and `bp-med`), 7 binary variables (`female`, `cvd-hx-baseline`, `statin`, `aspirin`, `antiarrhythmic`, `anti-coag`, and `x4smoke`), and 1 categorical variable (`raceclass`). These variables represent baseline demographic, biochemical, cardiovascular-risk, and treatment-related measures used to assess heterogeneous effects of intensive interventions on cardiovascular outcomes. The treatment variable (`treatment`) denotes intensive versus standard blood-pressure control in ACCORD-BP and intensive versus standard glycemic control in ACCORD-Glycemia. In both datasets, the outcome variable (`censor-po`) represents the primary composite cardiovascular event. Further details on inclusion criteria, participant demographics, variable definitions, feature distributions, and trial methodology are provided in the original ACCORD studies ^20**?**^ . For ACCORD and SPRINT, both datasets are available at BioLINCC repository upon request.

### A.2 Observational Data

#### Harborview pre-hospital TXA cohort

The emergency medicine datasets used in this study were gathered over 13 years (2007-2020) and encompass 14,463 emergency department admissions. It is the only retrospective electronic health record (EHR) data that we used in our study. We excluded patients under the age of 18 and patients with hypotension, and we curated a clinical cohort with patients prescribed tranexamic acid (TXA) among trauma patients and the corresponding control group. The corresponding cohort group is selected based on propensity score matching ^41^. The cohort consists of 240 patients with 120 in the treated group and 120 in the control group. We selected variables similar to CRASH-2 settings, including trauma type, demographic information (age, sex), and pre-hospital vital signs (blood pressure, heart rate, respiratory rate). The outcome was each patient’s survival. This dataset is not publicly available due to patient privacy concerns.

### A.3 Data Preprocessing

For our analysis, we curated each dataset by excluding records with substantial missingness (greater than 90%). For variables with missingness below this threshold, missing values were imputed using the empirical mean via the `SimpleImputer` function from the *scikit-learn* library. All continuous features were then normalized to the range [0, 1] using min–max scaling to ensure comparability across variables.

We also aggregated and removed redundant or overlapping features to simplify the feature space based on consultation with licensed physicians. For example, the Glasgow Coma Scale (GCS) for eye, motor, and verbal responses were combined into a single total GCS score. Additionally, highly correlated variables, such as the NIHSS score and its predicted counterpart, or overlapping indicators of stroke presence and symptom severity, were excluded.

### A.4 Semi-synthetic Data

In our synthetic data environment, we replicate the data generation process described in ^38^. This approach generates semi-synthetic data based on the Twins, News, and ACIC2016 datasets. It’s important to note that our primary focus is on varying confounding settings. Within these settings, treatment assignment is determined by:

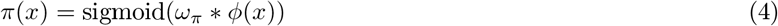

In this equation, *ω*_*π*_ stands for the confounding (propensity) scale, and *ϕ*(*x*) represents the confounding function. The confounding is either predictive, with *x* = *I*_*pred*_, prognostic, defined by *x* = *I*_*prog*_, or irrelevant, where *x* = *I*_*irrelevant*_.

#### Twins

The Twins dataset ^60^ encompasses 11,400 twin births in the USA from 1989 to 1991. It includes 39 covariates—both continuous and categorical, related to parents, pregnancy, and birth details.

#### News

The News dataset comprises 10,000 randomly sampled news items, each characterized by 2,858-word counts ^61,62^. The dataset is processed with Principal Component Analysis, with the first 100 principal components serving as continuous covariates for each item.

#### ACIC2016

The ACIC2016 dataset consists of data from the Collaborative Perinatal Project provided as part of the Atlantic Causal Inference Competition (ACIC2016) ^63^. It consists of 55 mixed (continuous and categorical) features for 2,200 patients.

## B Potential Outcome Framework

Using the Neyman–Rubin potential outcome framework ^37^, suppose a superpopulation *P* gives rise to a sample of *N* independent random variables, represented as (*Y*_*i*_(0), *Y*_*i*_(1), *X*_*i*_, *W*_*i*_) ∼ *P*. Here: - *X*_*i*_ ∈ ℝ^*d*^ denotes a *d*-dimensional feature vector. - *W*_*i*_ ∈ { 0, 1} signifies the treatment assignment, with the specific meaning to be clarified later. - *Y*_*i*_(0) and *Y*_*i*_(1) are the potential outcomes for unit *i* when assigned to the control and treatment groups, respectively. From this, we denote the Average Treatment Effect (ATE), as:

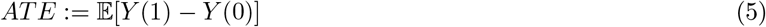

A core challenge in causal inference is the inability to observe both potential outcomes for each unit. For each unit, either the outcome under control (*W*_*i*_ = 0) or under treatment (*W*_*i*_ = 1) is observed, but not both. Given this, the observed data is:

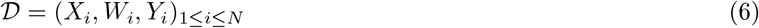

To decide the treatment for a new individual *i* with covariate *x*_*i*_, we aim to estimate its Individual Treatment Effect (ITE), *D*_*i*_ = *Y*_*i*_(1) − *Y*_*i*_(0). However, since *D*_*i*_ remains unobserved and is not identifiable without robust assumptions, we focus on estimating the Conditional Average Treatment Effect (CATE), *τ* (*x*):

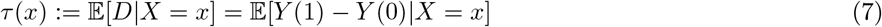

Note that the optimal estimator for CATE is also the best for ITE in terms of Mean Squared Error (MSE) ^2^. We aim for estimators that minimize the Expected Mean Squared Error (EMSE) for CATE estimation. This metric is also called the *precision in estimating heterogeneous effects (PEHE)*. ^64^.

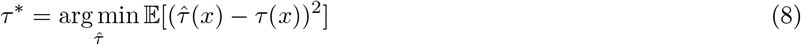

To be able to identify the causal effects from observational data, we make the standard assumptions for both domains.

### Assumption 1.

*(Unconfoundedness)* There are no unobserved confounders, such that the treatment assignment and POs are conditionally independent given the covariates:

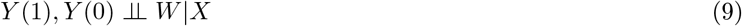

### Assumption 2.

*(Overlap)* for all *x* ∈ ℝ^*d*^,

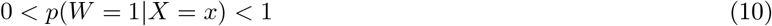

### B.1 CATE Models

#### B.1.1 Meta Learners

In this section, we provide the background of meta-learners mentioned in the main text.

**S-learner (Single Learner)** The S-learner trains a single model on both treated and control units.

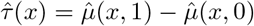

where 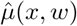 is the prediction of the model given covariate *x* and treatment *w*.

**T-learner (Two Learner)** ^2^ introduced the T-learner, an approach that creates distinct regression functions for each treatment group and computes the differences. The T-learner trains separate models for treated and control units.

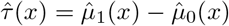

where 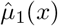 and 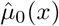 are the predictions for treated and control units, respectively.

**X-learner** ^2^ also introduce the X-learner, a two-step regression estimator that uses each data point twice. The X-learner is designed to leverage both the treated and control groups for estimating heterogeneous treatment effects. To start, models 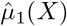 and 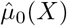 are fitted on the treated and control groups respectively. Using these models, treatment effects for the treated group are calculated as 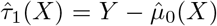 and for the control group as 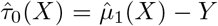. Subsequently, counterfactual outcomes for the treated group are estimated using *g*_0_ to get 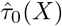 and for the control group using *g*_1_ to get 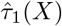. The final treatment effect estimate integrates these results using the propensity score, *e*(*X*) = *P* (*W* = 1|*X*)

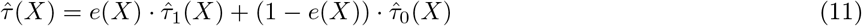

**DR-learner (Doubly Robust Learner)** ^42^ introduced the DR-learner, or doubly robust learner, a two-step procedure that employs the formula for the doubly robust augmented inverse propensity weighted (AIPW) estimator ^65^ as a pseudo-outcome in a two-step regression framework. The DR-learner combines propensity score weighting with regression adjustment.

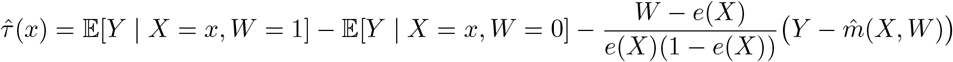

where *e*(*X*) is the propensity score.

**R-learner** ^66^ proposed the R-learner, which demands an estimate of the treatment-unconditional mean. The R-learner utilizes residuals to estimate the heterogeneous causal effect. Given a model, the residuals are computed as 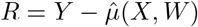. The causal effect is then estimated as:

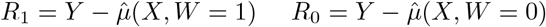

The causal effect is deduced from the relation between the residuals and the treatment assignment, given by:

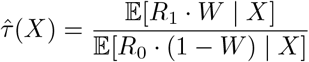

In this approach, the essence is to determine how deviations from the model’s predictions (residuals) correlate with the treatment, adjusted by the propensity of receiving the treatment.

#### B.1.2 Representation Learners

In addition to meta-learners, recent studies aim to learn a covariate shift function ^61^, ensuring that the feature representation of both treated and untreated groups have similar distributions. Algorithms such as CFRNet ^61^, TARNet ^67^, and Dragonnet ^68^ are introduced to predict CATE using balanced representations derived from observational data.

Specifically, the balanced learning approach identifies a representation, *h* : *X* → ℛ, and treatment-specific functions, *µ*_1_ and *µ*_0_, which aim to minimize the PEHE evaluation measure. The model is trained using a loss function that’s bounded by PEHE. This function comprises the combined expected factual treated and control losses for outcome regression and the distance between *h*(*x*) for given *W* = 1 and *W* = 0 values concerning the covariate shift.

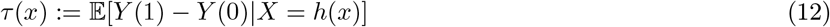

##### Dragonnet

Dragonnet employs a neural network architecture where both the treatment assignment and the outcome are jointly modeled. The network is structured to provide a representation *h*(*X*) of the input covariates that captures the nuances needed to predict both treatment propensity and potential outcomes. By jointly modeling, Dragonnet ensures that the learned representations are informative about both treatment assignments and outcomes.

##### TARNet (Treatment Agnostic Representation Network)

TARNet aims to learn a shared representation *h*(*X*) of the covariates for both potential outcomes. Once this representation is determined, it is then passed through two separate outcome models to predict the potential outcomes under treatment and control:

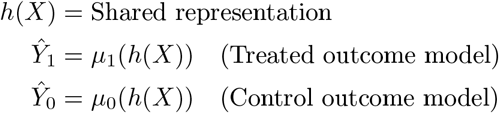

The shared representation ensures that the outcome predictions are based on a consistent understanding of the covariates.

##### CFR (Counterfactual Regression)

CFR focuses on deriving a shared representation *h*(*X*) that can produce balanced representations of treated and control units. This balance ensures that the distributions of the treated and control units in the representation space are similar, minimizing the distributional shift and aiding in counterfactual prediction. With this balanced representation, potential outcomes are then estimated using respective outcome models.

##### DR-CFR (Doubly Robust Counterfactual Regression)

DR-CFR combines the strengths of doubly robust estimation methods with the representation learning of CFR. After learning a balanced representation *h*(*X*), DR-CFR not only predicts potential outcomes but also adjusts for discrepancies using propensity scores or outcome residuals, leading to a more robust and accurate estimate of treatment effects.

### B.2 Model Implementation & Hyperparameters

For CATE model implementations, for S-Learner, T-Learner, X-learner, DR-learner, Dragonnet, TARNet, CFR, DR-CFR, we rely on `CATENets` package^3^. For CausalForest and Linear DR-Learner, we follow the implementation from ^4^. Each nuisance function (i.e., 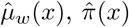, and 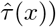 was constructed with the same number of hidden layers and hidden units across models. For architectures such as TARNet and CFRNet, the representation network Φ and the outcome heads *h*_*w*_ each contained one hidden layer with dense units and ReLU activations. For hyperparameter selection, we performed a grid search over the following ranges: training epochs, learning rates, batch sizes, numbers of hidden layers, and hidden dimensions. All models were trained using the Adam optimizer with early stopping based on validation performance.

**Table S.1.** Hyperparameter ranges for CATE models.

| Hyperparameter | Range |
| --- | --- |
| Training epochs | {10, 20, 30, 40, 50, 80, 100} |
| Learning rate | {$10^{-2}$, $10^{-3}$, $10^{-4}$, $10^{-5}$} |
| Batch size | {32, 64, 128} |
| Number of hidden layers | {1, 2, 4} |
| Hidden dimensions | {128, 256, 512} |
| Optimizer | Adam |
| Regularization constant (CFR) | {0, $10^{-1}$, $10^{-2}$, $10^{-3}$} |

### B.3 CATE Model Evaluation

Here we introduce different evaluation strategies to validate CATE models.

**Factual criteria**. ^47^ evaluate models by measuring simple prediction loss only on the observed potential outcomes.

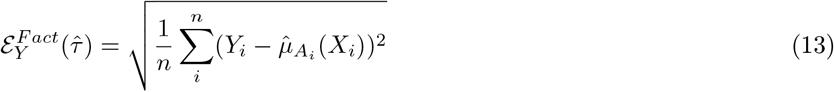

Obviously, one obvious disadvantage is that it may wrongly prioritize good fit on the potential outcomes over good CATE fit, resulting in bias ^47^.

**Plug-in criteria**. construct surrogates for CATE evaluation by fitting a new CATE estimator 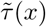 on held-out data and using this to compare against the estimates.

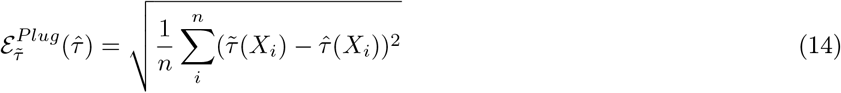

However, one obvious disadvantage for a plug-in surrogate is that it would favor models with similar structures e.g. S-Learner and T-Learner, a phenomenon called congenital bias. ^38,47^

**Pseudo-outcome surrogate criteria** obtain estimate through given auxiliary nuisance estimates 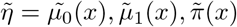 obtained from the validation data using ML method M, one can construct pseudo-outcomes 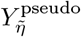 for which it holds that for ground truth nuisance parameter *η*, E[*Y*_*η*_ | *X* = *x*] = *τ* (*x*) and – instead of using them as regression outcomes as in the learners themselves

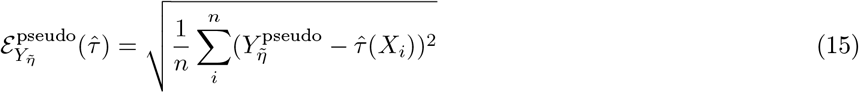

Here 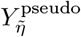 can be any pseudo-outcome objectives. In this work, we employ *Influence function, DR-Learner* objective, and *R-Learner* objective. Proposed by ^42^, DR pseudo-outcome employs doubly robust AIPW estimator and is hence unbiased if either propensity, 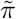, or outcome regressions, *ũ*_1_ and *ũ*_0_, are correctly specified. 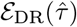 and 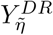 are denoted as,

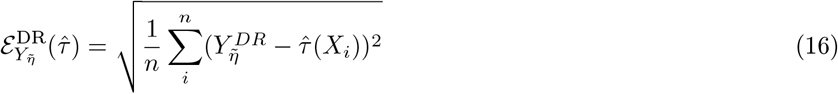

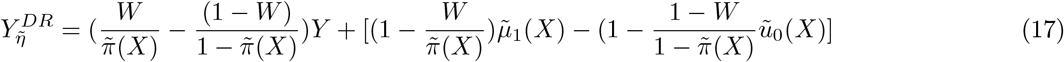

The R-learner objective of ^66^, which requires an estimate of the treatment-unconditional mean *µ*(*x*) = *E*[*Y* | *X* = *x*], relies on a similar idea and can also be used for the selection task, resulting in the criterion.

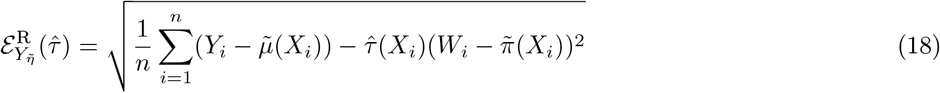

The influence function objective ^46^, categorized as surrogate pseudo-outcome ^47^, leverages Talyor-like expansion to approximate the PEHE.

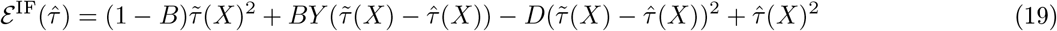

where 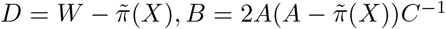 and 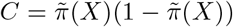, and 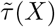 is a plug-in estimate. Surrogate pseudo-outcome is an unbiased estimator for CATE and more robust to congenital bias ^47^. On the theoretical side, the analysis in ^69^ shows that – under certain technical conditions – minimizing the surrogate validation estimate is a consistent model selection rule.

#### Qini Score and Uplift score

While pseudo-outcome surrogate provides an estimation for model selection, evaluating the performance of the chosen model on a different cohort can be difficult. As it requires training a second CATE model and nuisance functions. Therefore, the Qini curve is often used to evaluate model performance across different cohorts. It is defined as follows:

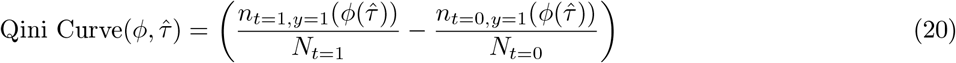

where 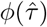 is the fraction of the population treated (in either treatment *t* = 1 or control *t* = 0 ) ordered by the uplift (treatment effect) from the model, 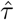. *N*_*t*_ represent the total individual count in each group. Similarly, the uplift score can also be used to evaluate CATE model but only considering the treatment uplift within *ϕ* population. The uplift score is calculated as:

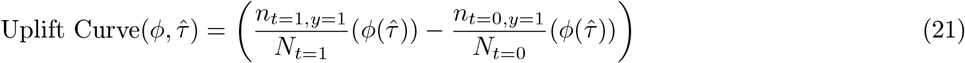

A baseline method is a model that cannot distinguish positive and negative uplift, and rank patients at random^5^. To quantify uplift/treatment effect from a given model, it is common to measure the area under the curve (AUROC) between the Qini/Uplift curve (intervention assignment based on model predictions) and random choice.

## C Interpretability (XAI) Methods

### C.1 Key Explanation Properties for CATE Models

Considering the CATE setting, ^38^ suggests the following key properties for explanation methods, *a*_*i*_, *i* ∈ [*d*], in a CATE model.

#### Sensitivity

The covariates that do not affect the CATE model are given zero contribution. More formally, if for some *i* ∈ [*d*] we have 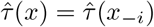 for all *x* ∈ *X*, then 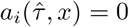 for all *x* ∈ *X* .

#### Completeness

Summing the importance scores gives the shift between the CATE and a baseline. More formally, for all *x* ∈ *X*, we have:

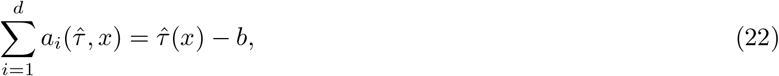

where *b* ∈ *R* is a constant baseline. In this way, each importance score *a*_*i*_ can be interpreted as the contribution from covariate *i* of*x* to have a CATE that differs from the baseline *b*. Note that the choice of the baseline differs from one method to another. For instance, the baseline for SHAP ^7^ is the average treatment effect: 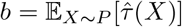.

#### Linearity

The importance score of a covariate is linear with respect to a black-box function. If the CATE model 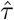 is written in terms of the estimated potential outcomes 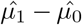, it can be written as 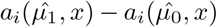. This makes it easy to differentiate prognostic and predictive covariates. If *x*_*i*_ is a prognostic covariate, then 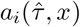 is zero. If *x*_*i*_ is a predictive covariate, then 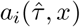 is not zero.

#### Model Agnosticism

The feature importance score can be computed for all CATE 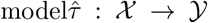. Some methods only work with a restricted family of models, which prevents them from being model agnostic. For example, Gradient-based explanation methods ^50,70^ only work for 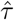 that is differentiable with respect to its input.

#### Implementation Invariance

The feature attribution would be the same for two functionally equivalent models. This means that if we have two CATE models 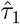 and 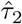 such that 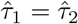 for all *x* ∈ *X*, this implies that 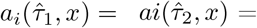 for all *x* ∈ *X* and all *i* ∈ [*d*].

### C.2 Explanation Methods

In this study, we utilize explanation methods that fulfill the following criteria, including *sensitivity, completeness, linearity*, and *implementation invariance*. Accordingly, we employ Integrated Gradients ^49^ and Shapley values ^7^. Although Vanilla Gradient (Saliency) ^50^ doesn’t satisfy the completeness criterion, it’s served as a basic baseline. All the feature importance methods are implemented using Pytorch and Captum Python package^6^.

#### Vanilla Gradient

The vanilla gradient, often referred to as the saliency map ^50^, is frequently used as a benchmark for different explanation techniques. It is derived from the gradient of the output function in relation to the input features. Specifically, within CATE models, it assesses how the estimated treatment effect changes with respect to a given feature *x*_*i*_.

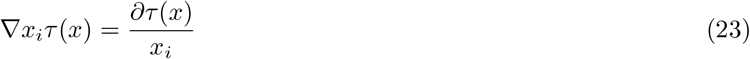

#### Integrated Gradients

Integrated Gradients (IG) assigns importance to input features by approximating the integral of a model’s gradients from a baseline input to the actual input ^49^. This method provides a holistic view of feature contributions, satisfying key properties like completeness. The IG attribution for an explicand *x*, a variable *x*_*i*_, and a baseline *x*^′^ is:

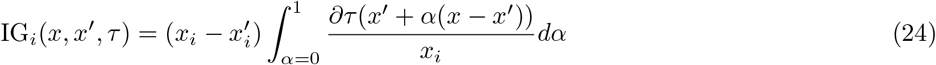

We use the zero vector as the baseline, denoted as *x*^′^ = 0. This means feature contributions are measured relative to their absence.

#### Shapley Value Sampling

Shapley Value, a concept borrowed from cooperative game theory, offers a unique approach to feature attributions ^7^. For any prediction model, it assigns each feature an importance value by averaging all possible combinations of feature presence or absence. Mathematically, for a prediction model *τ*, the exact Shapley value for a feature *x*_*i*_ is defined as:

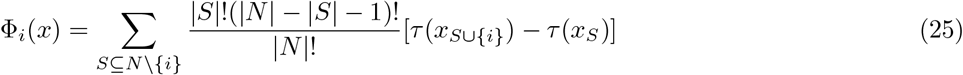

Where *N* is the set of all features and *S* is any subset of *N* that does not include feature *x*_*i*_. This equation evaluates the contribution of the feature *x*_*i*_ by contrasting the prediction with and without the feature over all possible combinations.

However, computing the exact Shapley value can be computationally intensive, especially for models with a large number of features. Therefore, in practice, an approximation method like Shapley Value Sampling ^71^ is often used.

Given a feature set *N*, and for a particular feature *x*_*i*_, the sampled Shapley value is estimated as:

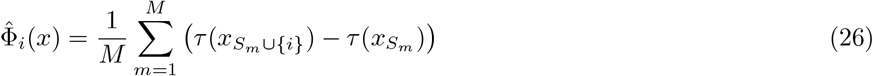

Where *M* is the number of sampled orderings and *S*_*m*_ is a random subset of *N* without feature *x*_*i*_ in the *m*^*th*^ sampled ordering. By sampling, we approximate the Shapley value, making it feasible for practical applications while maintaining a close approximation to the exact value.

#### Baseline Shapley

To obtain explanation/feature attribution for CATE models, we used the Shapley value ^6,7^, which is the average expected marginal contribution of adding one feature to the treatment effect after all possible combinations of features have been considered. More formally, the Shapley value takes as input a set function *v* : 2^*N*^ → *R*. The Shapley value produces attributions *s*_*i*_ for each player *i* ∈ *N* that add up to *v*(*N* ). The Shapley value of a player *i* is given by:

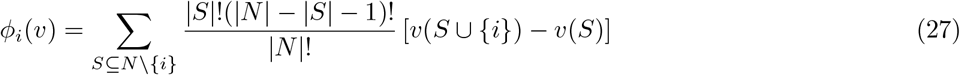

In other words, the Shapley value of player *i* is the average weighted average of its marginal contribution *v*(*S* ∪ {*i*} ) − *v*(*S*). Since computing the exact shapley value is intractable, several approximation methods have been proposed ^7,71^. Also, different choices of set function could lead to different Shapley values *ϕ*(*i*). One choice is to define *v*_*f,x*_(*S*) as the conditional expected model output on a data point when only the features in *S* are known ^45^

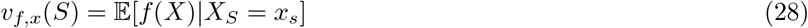

However, computing conditional distribution, 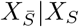, is usually difficult. Empirically, methods such as KernelSHAP ^7^ and shapley value sampling ^71^, conditional expectations are estimated by assuming feature independence; samples of the features in 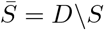 are drawn from the marginal joint distribution of these variables.

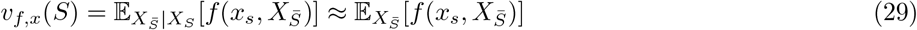

Moreover, we also leverage the assumption of model linearity to simplify the computation of the expected values ^7^. Empirically, we approximate the marginal distribution with an empirical mean of features as our baseline.

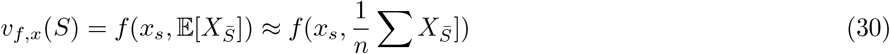

This approach is called Baseline Shapley (BShap) ^45^ and is more computationally efficient. More generally, this approach models a feature’s absence using its value in the baseline *x*^′^.

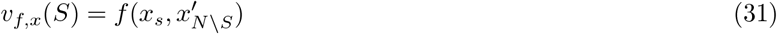

Another extension from baseline Shapley is Random Baseline Shapley (RBShap) ^45,72^ where the baseline 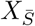 is drawn randomly according to the data distribution *D*. When propagating Shapley values, it approximates equation 29. However, we observed that this approach requires a significant amount of time to reach convergence. Therefore, in this work, we utilize BShap to explain CATE models.

#### Implementation details

For post-hoc interpretability at the individual level, we compared Shapley values with alternative attribution methods, including Integrated Gradients (IG) ^49^, gradient-based Saliency maps ^50^, Smooth-Grad ^51^, and LIME ^52^. All methods were implemented in `Captum` (v0.6.0) with respect to the CATE prediction 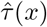. For Saliency, we used the `Saliency` class without smoothing; for IG, a mean baseline and 50 steps; for SmoothGrad, 50 noisy samples with Gaussian perturbations (*σ* = 0.2) averaged with `NoiseTunnel`; for Shapley, 500 permutations with the mean input as baseline; and for LIME, 500 perturbed samples with kernel width 0.75.

To obtain global feature rankings, we aggregated local attributions by taking the mean of their absolute values across individuals. For ensembles, attributions were computed for each model separately and then averaged across models.

### C.3 Examination of Explanation Method in CATE

#### Predictive Features Identification with Ground Truth

Following the works of ^38,44^, within a semi-synthetic environment, the health outcome, denoted as *µ*_*W*_, can be expressed as a function of the patient characteristics *X* and the treatment *W* :

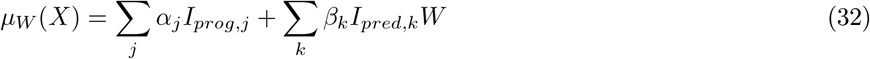

Where *I*_*prog,j*_ and *I*_*pred,k*_ denote the *j*^*th*^ and *k*^*th*^component of *X* that are prognostic and predictive covariates respectively. *α*_*j*_ and *β*_*k*_ are the corresponding coefficients. Under this assumption, the treatment effect can then be defined as:

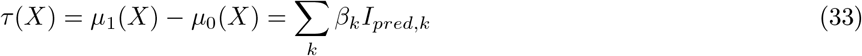

Therefore, in the context of CATE estimation, an explanation method should differentiate between predictive and prognostic covariates. To evaluate this, we can compute the average proportion of attribution correctly assigned to the predictive covariates:

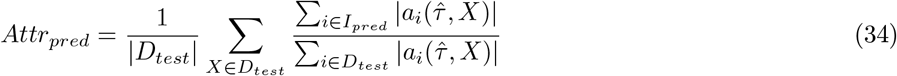

In this equation, *a*_*i*_ denotes the attribution score for a given feature *i*. This metric principally assesses the ability of an explanation method to differentiate between predictive and prognostic factors. While actual explanations or ground truths are absent in counterfactual predictions, this approach can offer guidance when choosing which explanation method is best suited for real-world datasets.

#### Ablation and Insertion and Deletion with Pseudo-outcomes

In practical scenarios, it’s usually impractical to obtain oracle features, therefore, to overcome this, one way is to perform an ablation test with CATE models. The ablation test is a commonly used evaluation technique for attribution values ^56^. Its concept involves replacing features with baseline feature values based on their attributions to measure their impact on the evaluation metric. This can be iteratively described using modified versions of the original explicands. For a given mixture of explicands, denoted by 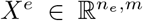, the attribution score is represented as *ϕ*(*f, X*^*e*^). The ablation study is characterized by three distinct parameters: (1) feature ordering, (2) an imputation sample, represented as *x*^*b*^ ∈ ℝ^*m*^, and (3) an evaluation metrics.

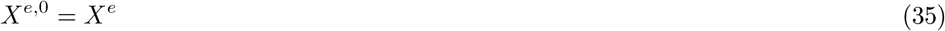

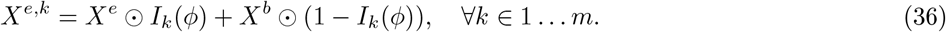

Where *X*^*b*^ := [*x*^*b*^ … *x*^*b*^]^*T*^ and *I*_*k*_(*ϕ*(*f, X*^*e*^)) = arg max_*k*,axis=1_(*ϕ*(*f, X*^*e*^)). The latter yields an indicator matrix the same size as *G*. A value of 1 indicates its position among the maximum *k* elements across a specific axis. The operation ⊙ represents the Hadamard product. Further, to assess the results of the ablation test, we compute the mean CATE output. When *k* features are ablated, this average is given by:

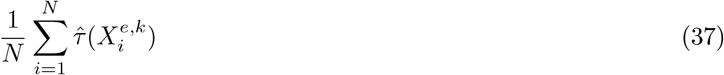

To contextualize the ablation test using treatment effects, imagine these features represent different features in a clinical trial, and their attributions (*ϕ* indicate the importance of each feature in predicting a particular effect of treatment, say patient recovery. When employing ablation: (1) Positive ablation: If we ablate (remove) features with the most positive attributions (those believed to have the most significant positive effect), we’d anticipate a substantial drop in, for example, patient recovery rates (mean model output). As we progressively remove additional features, this rate will continue to decline but at a diminishing pace. In this scenario, steeper declines imply that our attributions of features’ contributions are accurate. (2) Negative ablation: on the other hand, when removing features with the most negative attributions (those believed to hinder recovery), we’d expect recovery rates to increase significantly. More steep inclines would indicate better attributions, suggesting that the dropped features were indeed important.

In addition to CATE output, we can also leverage pseudo-outcome surrogates Section B.3, measuring the estimated PEHE when features are removed.

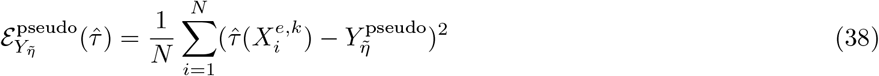

The term 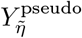 refers to the pseudo-outcome, such as the R or DR objectives, further detailed in Section B.3.

In addition to analyzing positive or negative ablations, insertion and deletion tests provide another way to assess performance. To conduct an insertion test, start with a baseline where no features are present and then gradually add features based on their ranking. Similarly, when assessing recovery rates, positive attributions should improve them while negative ones should have a negative impact.

#### Examining Interpretability Methods

Because ground-truth feature contributions to treatment effects are rarely observable, we evaluate explanation methods using indirect but complementary criteria: (i) *local fidelity*, which tests whether attributions reflect model behavior under perturbations; (ii) *global sufficiency*, which examines whether the most important features identified by the explanation are sufficient to recover model predictions; and (iii) *consistency*, which measures the stability of explanations across independently trained models. When external ground-truth signals are available, we additionally evaluate (iv) *feature rank concordance*.

##### Local fidelity

Local fidelity ^55^ measures how well feature attributions capture the model’s prediction behavior at the individual level. We adopt standard insertion-deletion benchmarks ^73^ with the pseudo–outcome surrogate (Section B.3) as the fidelity metric. This assesses whether removing or inserting features with high attribution values leads to the expected changes in predicted treatment effects.

##### Global sufficiency

Global sufficiency assesses whether features identified by an explanation capture the model’s predictive signal ^56^. We rank features by their attribution values,^7^ then train a student model 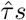 using only the top-*k* features to reproduce the ensemble’s predictions 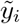.^8^ The distillation error over *M* test points is

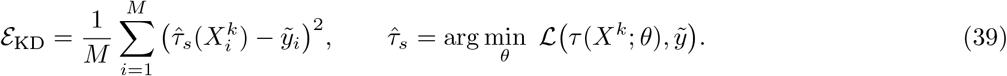

Lower ℰ_KD_ indicates that the explanation-selected features capture the teacher’s signal with fewer inputs.

##### Consistency

For explanations to be useful in scientific discovery, explanations must be stable across repeated training runs. We therefore assess consistency across *L* independently trained ensembles, 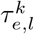 (with *L* = 1 corresponding to a single model), each comprising *k* base learners. We compute the mean pairwise cosine similarity of their *global* attribution vectors (average absolute attribution across individuals):

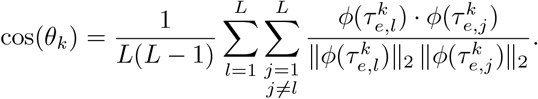

An increase in cos(*θ*_*k*_) with *k* indicates reduced sensitivity to random initialization and greater reflection of underlying signal rather than noise.

##### Feature rank concordance

When external ground-truth signals are available, we also assess rank concordance. Specifically, we compute *Spearman’s rank correlation* ^74^ between the global feature ranking from an explanation method, 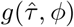, and oracle rankings *g*(*p*) derived from interaction *p*-values ^75^, where lower *p*-values denote stronger evidence of treatment effect heterogeneity. The resulting correlation

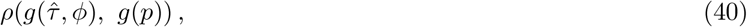

indicates whether explanation methods recover features consistent with ground truth features.

**Table S.2.** Model performance on IST-3, CRASH-2, SPRINT, and ACCORD. Metrics include pseudo-surrogate outcomes (ℰ_if-pehe_, ℰ_R_, ℰ_DR_) and incremental gain scores (Qini and Uplift). Results are reported as mean ± standard deviation across random seeds. Higher values indicate better performance for Qini and Uplift, whereas lower values indicate better performance for ℰ_R_, and ℰ_DR_.

|  | IST-3<sup>18</sup> |  | CRASH-2<sup>17</sup> |  | SPRINT<sup>21</sup> |  | ACCORD<sup>20</sup> |  |
| --- | --- | --- | --- | --- | --- | --- | --- | --- |
|  | DR-Learner | DR-Learner (Ens.) | DR-Learner | DR-Learner (Ens.) | DR-Learner | DR-Learner (Ens.) | RA-Learner | RA-Learner (Ens.) |
| Qini | 0.284 (0.034) | **0.358 (0.016)** | 0.136 (0.004) | **0.186 (0.004)** | 0.066 (0.003) | **0.117 (0.003)** | 0.096 (0.007) | **0.212 (0.008)** |
| Uplift | 0.300 (0.025) | **0.395 (0.019)** | 0.248 (0.005) | **0.316 (0.008)** | 0.125 (0.005) | **0.212 (0.005)** | 0.183 (0.012) | **0.358 (0.011)** |
| $\mathcal{E}_R$ | 0.521 (0.008) | **0.520 (0.008)** | **0.373 (0.003)** | 0.373 (0.003) | 0.241 (0.004) | **0.236 (0.006)** | 0.305 (0.005) | **0.300 (0.005)** |
| $\mathcal{E}_{DR}$ | 0.921 (0.016) | **0.919 (0.015)** | 0.684 (0.005) | **0.678 (0.004)** | **0.473 (0.007)** | 0.472 (0.010) | 0.592 (0.009) | **0.580 (0.009)** |

## D Additional Experiment Results

### D.1 CATE Evaluation in Clinical Datasets

In evaluating the Conditional Average Treatment Effect (CATE), we employed the pseudo-outcome surrogate approach, Section B.3. The experimental results are aggregated from 50 iterations of each model, each initialized with unique random seeds. A significant variance was observed with the influence function-based surrogate outcome metric, *ϵ*_if-pehe_. Owing to this instability, our model selection was principally driven by the more robust metrics, *ϵ*_*DR*_ and *ϵ*_*R*_, aligning with the findings of ^47^.

Our evaluations revealed minimal disparity in terms of the pseudo-outcome surrogate metrics *ϵ*_*DR*_ and *ϵ*_*R*_. Given that a majority of our datasets are sourced from randomized control trials—except the pre-hospital TXA dataset—this is anticipated. Especially in scenarios where the confounding (propensity) scale approaches 0, we observed a consistent identification of significant features across various explanatory methodologies, corroborated by the work of Crabbe et al. ^38^. We also observed that, for representation learners, the computational time for explanation is significantly longer than CATE models. Given these considerations, the X-Learner was predominantly selected as the model to be interpreted throughout our experiments.

### D.2 Evaluating Ensemble CATEs’ efficacy

The second criterion involved comparing the ensemble-derived average treatment effect (ATE) estimates with the reported ATE values from each trial. In well-controlled randomized trials, where randomization minimizes confounding, the average of CATE estimates should closely align with the ATE in the absence of significant effect modifiers ^1^. For IST-3 and CRASH-2, the CATE models produced estimates closely aligned with the reported outcomes ^17,18^ (see Table S.4). However, in the blood pressure control trials, i.e., SPRINT and ACCORD, the CATE models provided higher ATE estimates than those reported: 1.6% for SPRINT and 1.2% for ACCORD, compared to 0.54% and 0.22%, respectively. Moreover, the analysis reveals that SPRINT, which demonstrated the efficacy of intensive blood pressure control, and the ACCORD study, which showed no significant treatment effects, align with the reported findings. These findings show that ensemble CATE models capture treatment effects at the cohort level, particularly in trials with substantial treatment effects.

**Table S.3.** Model performance on IST-3, CRASH-2, SPRINT, and ACCORD. Metrics include pseudo-surrogate outcomes (ℰ_if-pehe_, ℰ_R_, ℰ_DR_) and incremental gain scores (Qini and Uplift). Results are reported as mean ± standard deviation across random seeds. Higher values indicate better performance for Qini and Uplift, whereas lower values indicate better performance for ℰ_if-pehe_, ℰ_R_, and ℰ_DR_.

| Metric | XLearner | DRLearner | SLearner | RALearner | RLearner | TLearner | TARNet | DragonNet | CFRNet | CausalForest | LinearDR |
| --- | --- | --- | --- | --- | --- | --- | --- | --- | --- | --- | --- |
| IST-3 |  |  |  |  |  |  |  |  |  |  |  |
| Qini | 0.191 ± 0.023 | **0.284 ± 0.034** | 0.108 ± 0.023 | 0.182 ± 0.020 | 0.149 ± 0.017 | 0.169 ± 0.023 | 0.009 ± 0.019 | 0.009 ± 0.026 | -0.001 ± 0.013 | 0.214 ± 0.008 | 0.043 ± 0.013 |
| Uplift | 0.203 ± 0.025 | **0.300 ± 0.037** | 0.116 ± 0.024 | 0.193 ± 0.020 | 0.157 ± 0.017 | 0.181 ± 0.024 | 0.009 ± 0.020 | 0.009 ± 0.028 | -0.001 ± 0.014 | 0.233 ± 0.011 | 0.045 ± 0.013 |
| $\epsilon_{\text{if-pehe}}$ | 154.7 ± 127.7 | 250.0 ± 136.0 | 101.0 ± 128.4 | 138.6 ± 120.7 | 168.7 ± 129.1 | 129.9 ± 134.2 | 94.1 ± 97.6 | 94.6 ± 95.6 | 93.1 ± 96.2 | **73.9 ± 112.4** | 88.1 ± 108.8 |
| $\epsilon_R$ | 0.523 ± 0.006 | **0.521 ± 0.008** | 0.531 ± 0.007 | 0.526 ± 0.007 | 0.527 ± 0.008 | 0.526 ± 0.007 | 0.534 ± 0.007 | 0.534 ± 0.008 | 0.534 ± 0.008 | 0.526 ± 0.008 | 0.533 ± 0.007 |
| $\epsilon_{\text{DR}}$ | 0.924 ± 0.012 | **0.921 ± 0.016** | 0.941 ± 0.014 | 0.929 ± 0.013 | 0.932 ± 0.014 | 0.931 ± 0.013 | 0.947 ± 0.013 | 0.947 ± 0.013 | 0.947 ± 0.013 | 0.930 ± 0.014 | 0.946 ± 0.013 |
| ACCORD |  |  |  |  |  |  |  |  |  |  |  |
| Qini | 0.088 ± 0.006 | 0.105 ± 0.007 | 0.056 ± 0.006 | **0.147 ± 0.006** | 0.078 ± 0.008 | 0.097 ± 0.008 | -0.002 ± 0.008 | 0.003 ± 0.009 | -0.001 ± 0.008 | 0.100 ± 0.007 | 0.025 ± 0.004 |
| Uplift | 0.166 ± 0.011 | 0.191 ± 0.012 | 0.106 ± 0.012 | 0.187 ± 0.013 | 0.148 ± 0.014 | 0.184 ± 0.015 | -0.003 ± 0.014 | 0.005 ± 0.016 | -0.002 ± 0.016 | **0.272 ± 0.009** | 0.049 ± 0.008 |
| $\epsilon_{\text{if-pehe}}$ | 34.0 ± 74.4 | 124.5 ± 69.0 | -24.1 ± 66.8 | -7.3 ± 67.6 | 52.9 ± 65.9 | 1.3 ± 66.1 | 23.2 ± 42.8 | 21.4 ± 45.5 | 23.3 ± 44.3 | **-68.9 ± 81.3** | -10.8 ± 60.5 |
| $\epsilon_R$ | 0.310 ± 0.005 | 0.307 ± 0.006 | 0.318 ± 0.005 | **0.305 ± 0.006** | 0.311 ± 0.007 | 0.307 ± 0.007 | 0.320 ± 0.006 | 0.320 ± 0.006 | 0.320 ± 0.006 | 0.313 ± 0.006 | 0.320 ± 0.006 |
| $\epsilon_{\text{DR}}$ | 0.601 ± 0.010 | 0.595 ± 0.011 | 0.618 ± 0.010 | **0.592 ± 0.012** | 0.603 ± 0.013 | 0.596 ± 0.012 | 0.624 ± 0.009 | 0.623 ± 0.010 | 0.623 ± 0.010 | 0.609 ± 0.010 | 0.622 ± 0.010 |
| SPRINT |  |  |  |  |  |  |  |  |  |  |  |
| Qini | 0.059 ± 0.005 | 0.066 ± 0.003 | 0.035 ± 0.004 | 0.056 ± 0.004 | 0.054 ± 0.005 | 0.064 ± 0.002 | -0.000 ± 0.006 | 0.001 ± 0.004 | 0.003 ± 0.004 | **0.101 ± 0.006** | 0.014 ± 0.002 |
| Uplift | 0.114 ± 0.009 | 0.125 ± 0.005 | 0.068 ± 0.007 | 0.107 ± 0.007 | 0.104 ± 0.010 | 0.124 ± 0.004 | -0.001 ± 0.011 | 0.001 ± 0.007 | 0.006 ± 0.008 | **0.192 ± 0.009** | 0.026 ± 0.004 |
| $\epsilon_{\text{if-pehe}}$ | -82.9 ± 84.3 | -26.0 ± 81.5 | -111.9 ± 91.4 | -107.4 ± 87.4 | -43.5 ± 95.3 | -83.3 ± 81.9 | 37.5 ± 59.8 | 36.1 ± 61.4 | 34.0 ± 59.8 | **-157.7 ± 89.6** | -59.8 ± 68.9 |
| $\epsilon_R$ | 0.243 ± 0.005 | **0.241 ± 0.004** | 0.249 ± 0.005 | 0.244 ± 0.005 | 0.244 ± 0.005 | 0.242 ± 0.005 | 0.251 ± 0.005 | 0.251 ± 0.005 | 0.251 ± 0.005 | 0.245 ± 0.005 | 0.250 ± 0.005 |
| $\epsilon_{\text{DR}}$ | 0.476 ± 0.007 | **0.473 ± 0.007** | 0.488 ± 0.008 | 0.478 ± 0.008 | 0.477 ± 0.008 | 0.474 ± 0.008 | 0.492 ± 0.008 | 0.492 ± 0.008 | 0.492 ± 0.008 | 0.479 ± 0.008 | 0.491 ± 0.008 |
| CRASH-2 |  |  |  |  |  |  |  |  |  |  |  |
| Qini | 0.067 ± 0.004 | **0.136 ± 0.004** | 0.041 ± 0.004 | 0.054 ± 0.004 | 0.053 ± 0.005 | 0.066 ± 0.005 | 0.001 ± 0.005 | -0.000 ± 0.004 | 0.004 ± 0.004 | 0.072 ± 0.007 | 0.011 ± 0.003 |
| Uplift | 0.124 ± 0.009 | **0.248 ± 0.005** | 0.076 ± 0.007 | 0.099 ± 0.007 | 0.098 ± 0.009 | 0.122 ± 0.009 | 0.003 ± 0.009 | -0.000 ± 0.008 | 0.008 ± 0.007 | 0.132 ± 0.013 | 0.020 ± 0.005 |
| $\epsilon_{\text{if-pehe}}$ | -42.9 ± 177.9 | 139.8 ± 213.9 | -85.5 ± 165.3 | -60.5 ± 186.6 | 27.1 ± 191.8 | -12.9 ± 162.1 | 87.9 ± 131.2 | 93.0 ± 128.1 | 91.4 ± 131.7 | **-122.6 ± 172.4** | -8.4 ± 149.6 |
| $\epsilon_R$ | 0.374 ± 0.003 | **0.373 ± 0.003** | 0.377 ± 0.003 | 0.375 ± 0.003 | 0.376 ± 0.003 | 0.374 ± 0.003 | 0.379 ± 0.003 | 0.379 ± 0.003 | 0.379 ± 0.003 | 0.372 ± 0.003 | 0.379 ± 0.003 |
| $\epsilon_{\text{DR}}$ | 0.686 ± 0.005 | **0.684 ± 0.005** | 0.694 ± 0.004 | 0.690 ± 0.005 | 0.690 ± 0.005 | 0.687 ± 0.004 | 0.698 ± 0.004 | 0.697 ± 0.004 | 0.697 ± 0.004 | 0.683 ± 0.004 | 0.697 ± 0.004 |

**Table S.4.** Comparison of predicted average treatment effect (ATE) estimates from CATE models (with 95% confidence intervals) and reported ATE values (primary outcome differences) across four clinical trials.

| Cohort | Predicted ATE (95% CI) | Reported ATE (%) |
| --- | --- | --- |
| CRASH-2 | 1.1 (0.2 - 1.9) | 1.5 |
| IST-3 | 2.0 (0.3 - 4.0) | 2.0 |
| SPRINT | 1.6 (0.8 - 2.4) | 0.54 |
| ACCORD | 1.2 (-0.3 - 2.4) | 0.22 |

### D.3 Insertion & Deletion Procedure with Pseudo-outcomes - Real-world Data

In this section, we evaluate explanation methods using standard *insertion* and *deletion* experiments (see Appendix C). Relevant features are extracted with respect to the pseudo-outcomes, *ϵ*_pseudo_. We compute the Area Under the Curve (AUC) using a random baseline and normalize it by the number of features for each dataset. Our results show that the Shapley value method achieves the highest normalized AUC in the ACCORD, SPRINT, and CRASH-2 datasets, indicating superior performance. In contrast, for the IST-3 dataset, LIME performs best in insertion, while the Shapley value performs best in deletion. We also find that using ℰ_*IF*_ tends to be relatively unstable, exhibiting high standard deviation, whereas ℰ_*DR*_ and ℰ_*R*_ are more consistent and stable across experiments.

**Table S.5.**
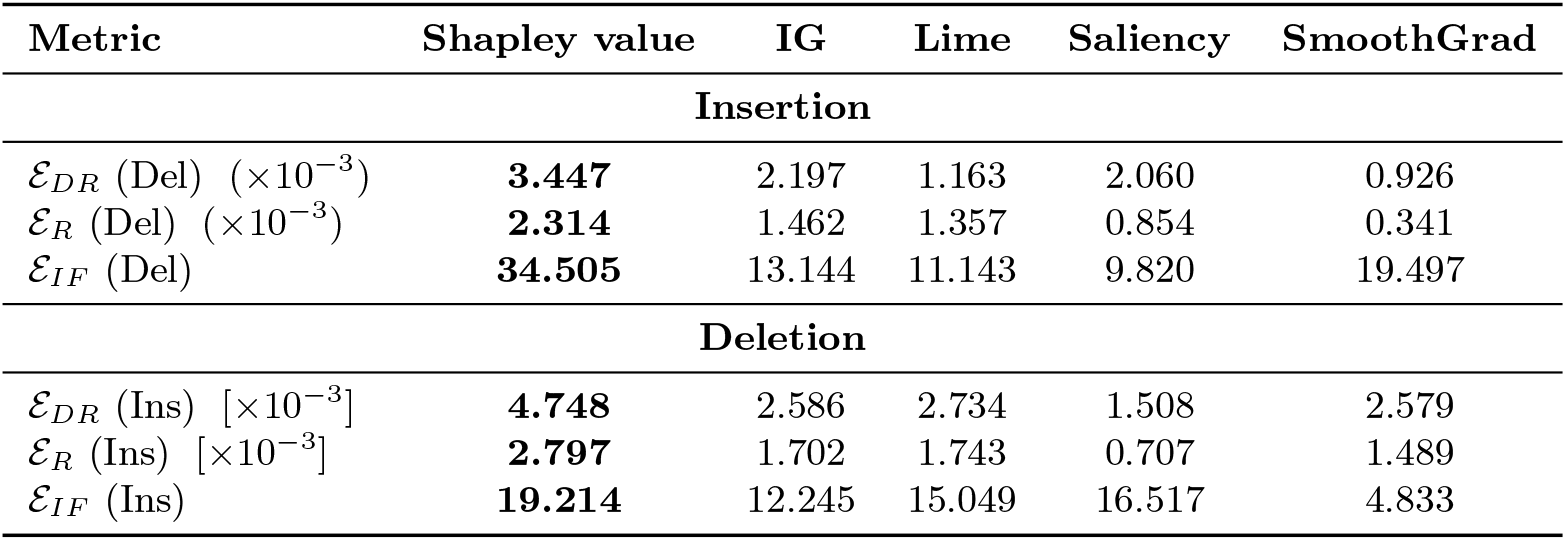
Normalized Area Under the Curve (AUC) for insertion and deletion curves on the ACCORD dataset. All values are normalized by the number of features. Results are reported for three pseudo-outcome surrogates: ℰ_*R*_, ℰ_*DR*_, and ℰ_*IF*_ . The best-performing method for each row is shown in **bold**.

| Metric | Shapley value | IG | Lime | Saliency | SmoothGrad |
| --- | --- | --- | --- | --- | --- |
| **Insertion** |  |  |  |  |  |
| $\mathcal{E}_{DR}$ (Del) ($\times 10^{-3}$) | **3.447** | 2.197 | 1.163 | 2.060 | 0.926 |
| $\mathcal{E}_R$ (Del) ($\times 10^{-3}$) | **2.314** | 1.462 | 1.357 | 0.854 | 0.341 |
| $\mathcal{E}_{IF}$ (Del) | **34.505** | 13.144 | 11.143 | 9.820 | 19.497 |
| **Deletion** |  |  |  |  |  |
| $\mathcal{E}_{DR}$ (Ins) [$\times 10^{-3}$] | **4.748** | 2.586 | 2.734 | 1.508 | 2.579 |
| $\mathcal{E}_R$ (Ins) [$\times 10^{-3}$] | **2.797** | 1.702 | 1.743 | 0.707 | 1.489 |
| $\mathcal{E}_{IF}$ (Ins) | **19.214** | 12.245 | 15.049 | 16.517 | 4.833 |

**Fig. S.1.**
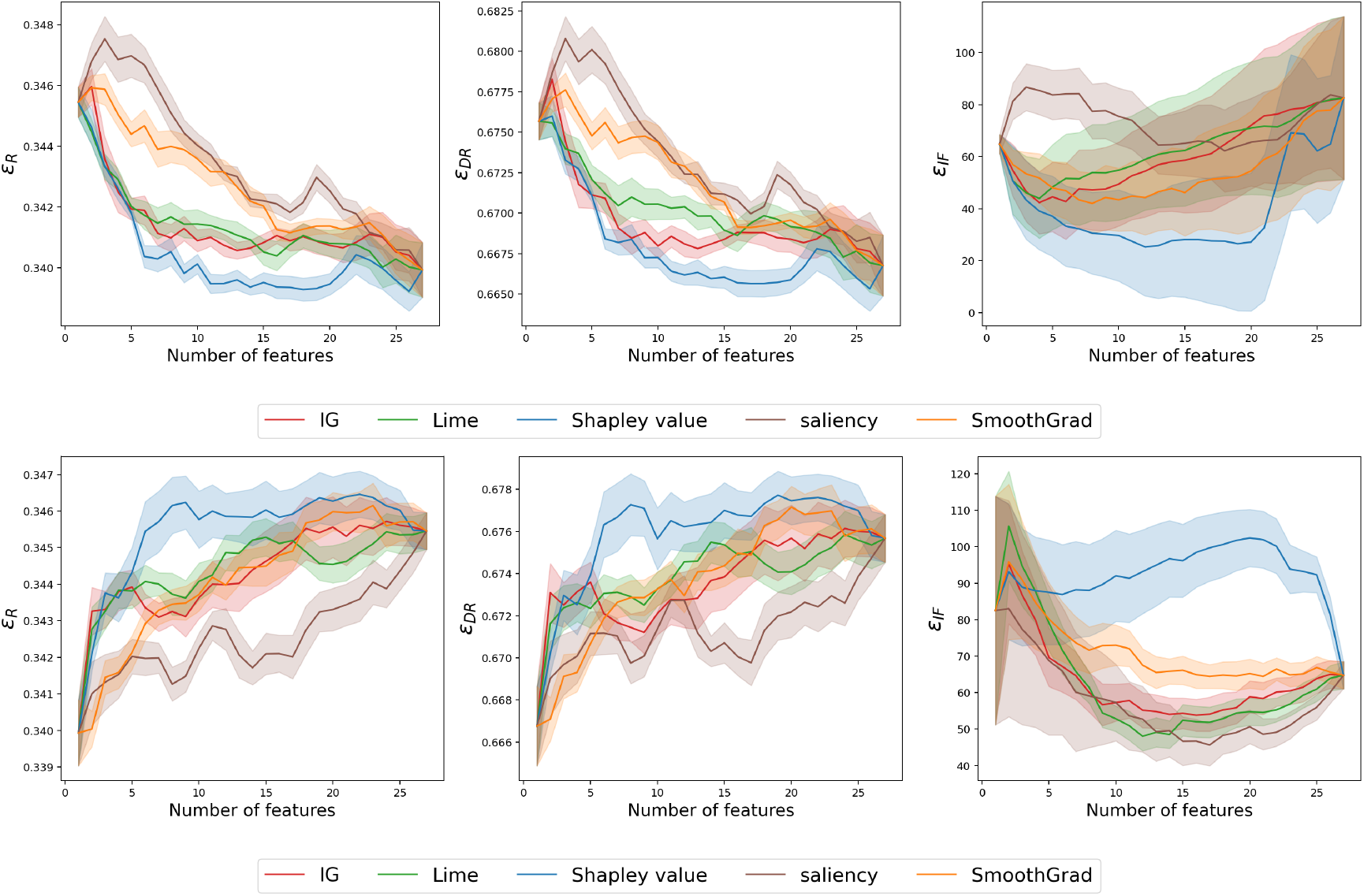
Insertion (top) and deletion (bottom) curves on the ACCORD dataset. The X-axis denotes the number of features removed or included, while the Y-axis shows the pseudo-outcome surrogates ℰ_*R*_, ℰ_*DR*_, and ℰ_*IF*_ .

**Table S.6.** Normalized Area Under the Curve (AUC) for insertion and deletion curves on the SPRINT dataset. All values are normalized by the number of features. Results are reported for three pseudo-outcome surrogates: ℰ_*R*_, ℰ_*DR*_, and ℰ_*IF*_ . The best-performing method for each row is shown in **bold**.

| Metric | Shapley value | IG | Lime | Saliency | SmoothGrad |
| --- | --- | --- | --- | --- | --- |
| **Insertion** |  |  |  |  |  |
| $\mathcal{E}_{DR}$ (Del) ($\times 10^{-3}$) | **2.573** | 1.294 | 1.725 | 1.079 | 1.140 |
| $\mathcal{E}_R$ (Del) ($\times 10^{-3}$) | **1.066** | 0.485 | 0.547 | 0.551 | 0.340 |
| $\mathcal{E}_{IF}$ (Del) | **33.271** | 22.262 | 27.084 | 8.440 | 8.914 |
| **Deletion** |  |  |  |  |  |
| $\mathcal{E}_{DR}$ (Ins) ($\times 10^{-3}$) | **2.289** | 1.356 | 1.228 | 0.591 | 0.937 |
| $\mathcal{E}_R$ (Ins) ($\times 10^{-3}$) | **1.044** | 0.776 | 0.583 | 0.272 | 0.497 |
| $\mathcal{E}_{IF}$ (Ins) | **51.544** | 38.708 | 47.688 | 12.237 | 29.173 |

**Fig. S.2.**
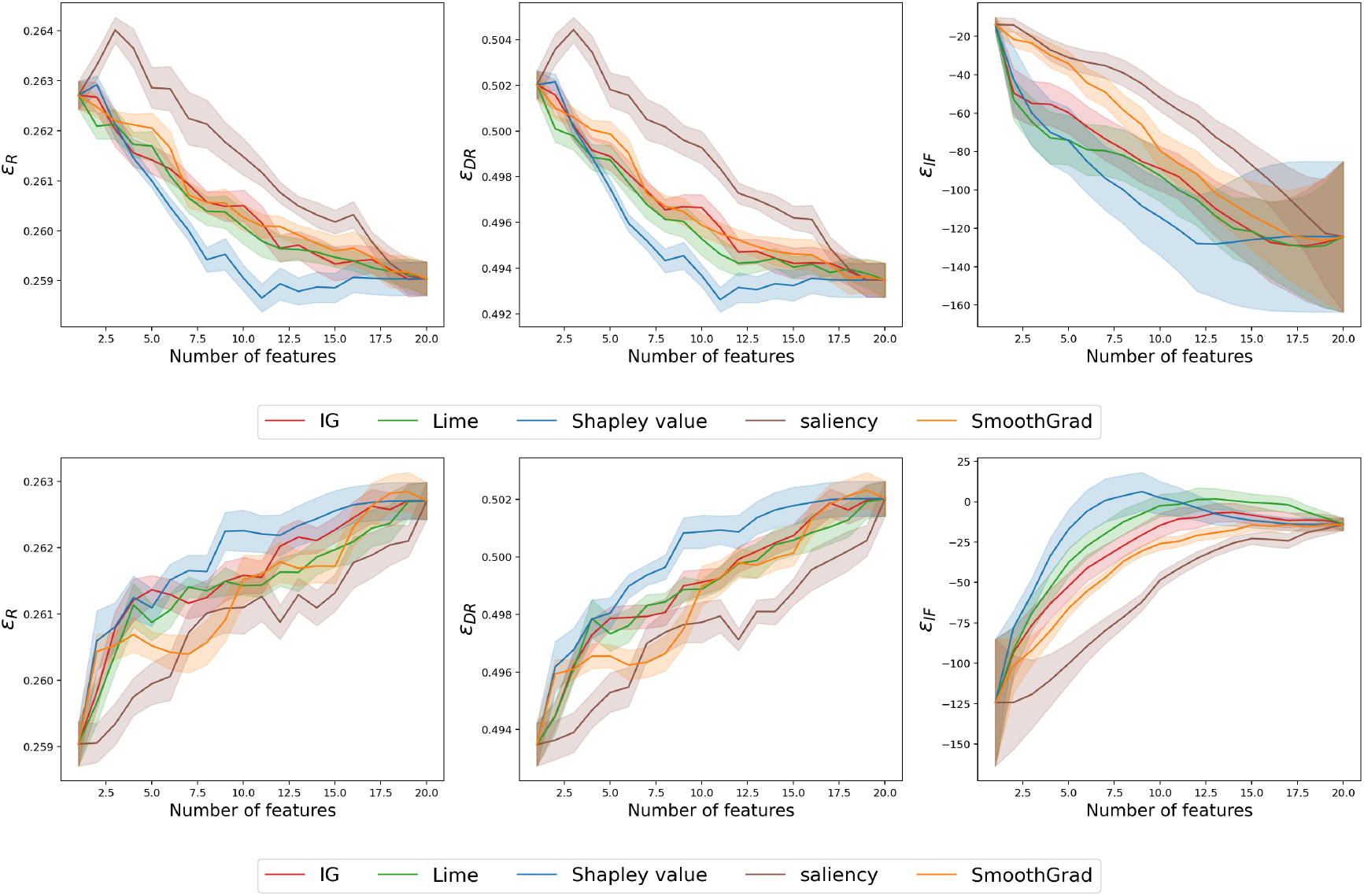
Insertion (top) and deletion (bottom) curves on the SPRINT dataset. The X-axis denotes the number of features removed or included, while the Y-axis shows the pseudo-outcome surrogates ℰ_*R*_, ℰ_*DR*_, and ℰ_*IF*_.

**Table S.7.**
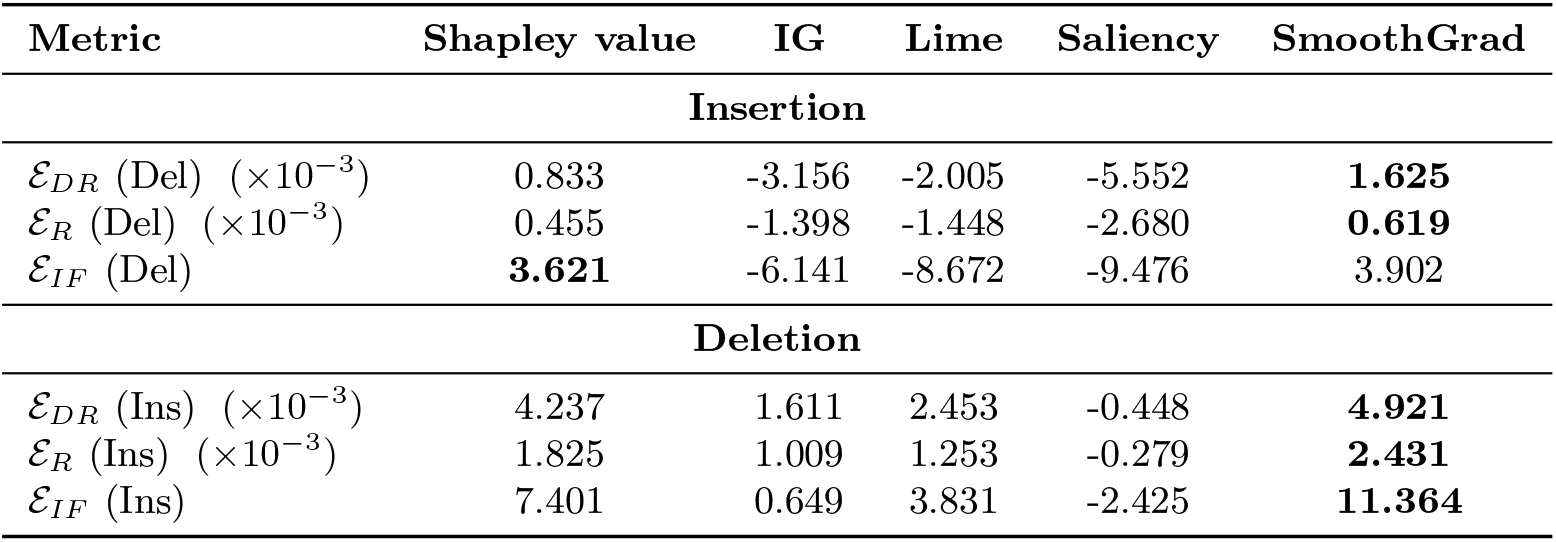
Normalized Area Under the Curve (AUC) for insertion and deletion curves on the IST-3 dataset. Value is normalized by the number of features. Results are reported for three pseudo-outcome surrogates: ℰ_*R*_, ℰ_*DR*_, and ℰ_*IF*_ . The best-performing method for each row is shown in **bold**.

| Metric | Shapley value | IG | Lime | Saliency | SmoothGrad |
| --- | --- | --- | --- | --- | --- |
| **Insertion** |  |  |  |  |  |
| $\mathcal{E}_{DR}$ (Del) ($\times 10^{-3}$) | 0.833 | -3.156 | -2.005 | -5.552 | **1.625** |
| $\mathcal{E}_R$ (Del) ($\times 10^{-3}$) | 0.455 | -1.398 | -1.448 | -2.680 | **0.619** |
| $\mathcal{E}_{IF}$ (Del) | **3.621** | -6.141 | -8.672 | -9.476 | 3.902 |
| **Deletion** |  |  |  |  |  |
| $\mathcal{E}_{DR}$ (Ins) ($\times 10^{-3}$) | 4.237 | 1.611 | 2.453 | -0.448 | **4.921** |
| $\mathcal{E}_R$ (Ins) ($\times 10^{-3}$) | 1.825 | 1.009 | 1.253 | -0.279 | **2.431** |
| $\mathcal{E}_{IF}$ (Ins) | 7.401 | 0.649 | 3.831 | -2.425 | **11.364** |

**Fig. S.3.**
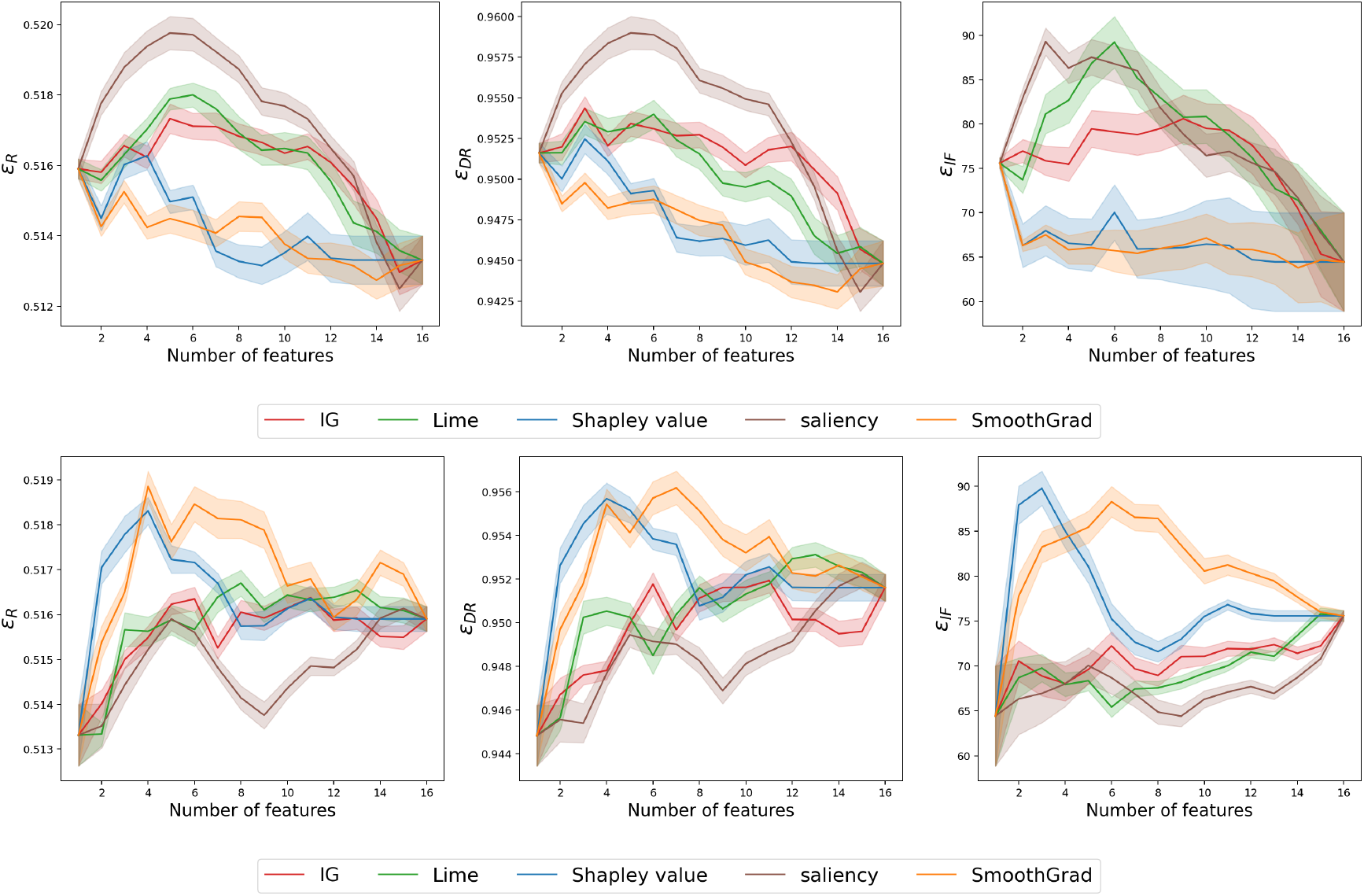
Insertion (top) and deletion (bottom) curves on the IST-3 dataset. The X-axis denotes the number of features removed or included, while the Y-axis shows the pseudo-outcome surrogates ℰ_*R*_, ℰ_*DR*_, and ℰ_*IF*_.

**Table S.8.**
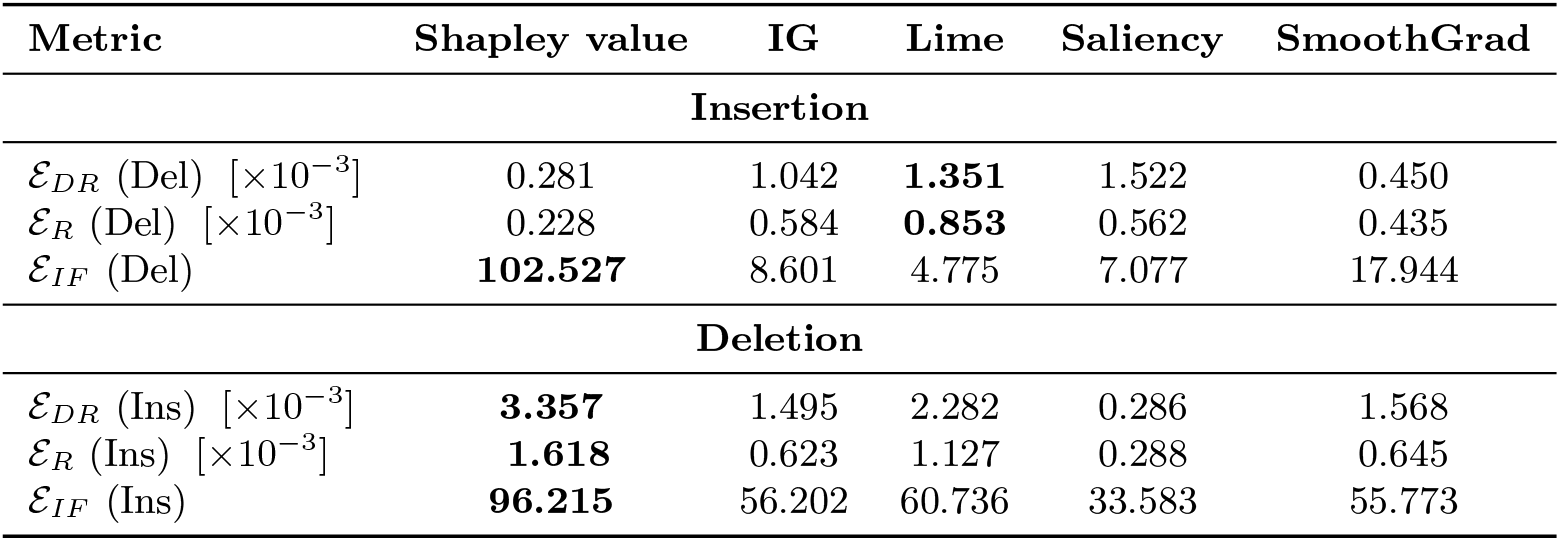
Normalized Area Under the Curve (AUC) for insertion and deletion curves on the CRASH-2 dataset. All values are normalized by the number of features. Results are reported for three pseudo-outcome surrogates: ℰ_*R*_, ℰ_*DR*_, and ℰ_*IF*_ . The best-performing method for each row is shown in **bold**.

**Fig. S.4.**
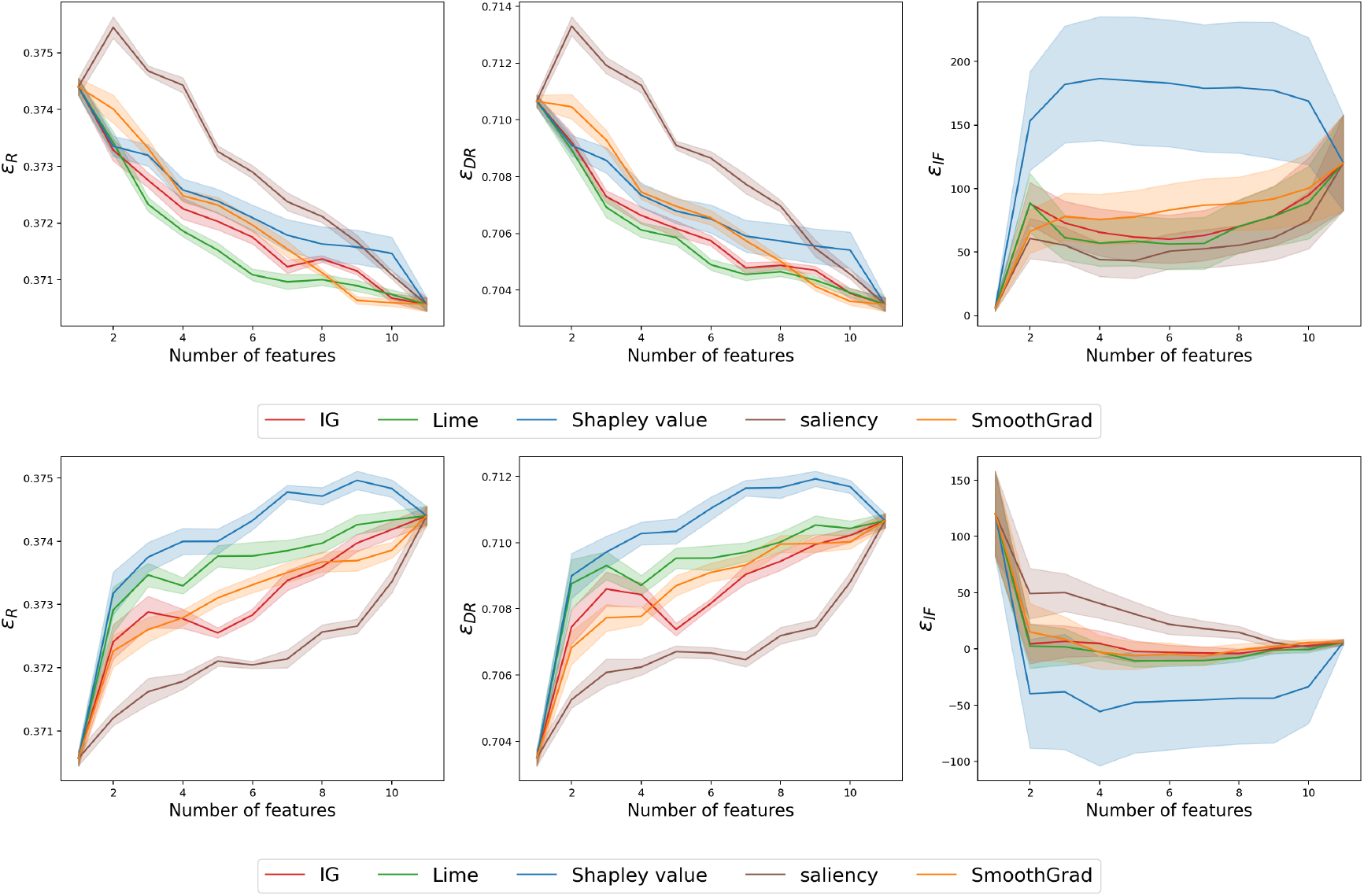
Insertion (top) and deletion (bottom) curves on the CRASH-2 dataset. The X-axis denotes the number of features removed or included, while the Y-axis shows the pseudo-outcome surrogates ℰ_*R*_, ℰ_*DR*_, and ℰ_*IF*_.

### D.4 Knowledge Distillation as a Benchmark Test in Clinical Datasets

**Table S.9.** Knowledge distillation performance and top 5 identified features for datasets IST-3, CRASH-2, SPRINT, and ACCORD. Black indicates the method with the lowest distillation loss with feature budgets at 5.

| Explanation Methods | MSE ($\times 10^{-3}$) | Top 5 Features |
| --- | --- | --- |
| CRASH-2 |  |  |
| Saliency | 4.0 ($\pm 0.2$) | heart rate, injury time, respiratory, capillary refill time, systolic blood pressure |
| SmoothGrad | 3.8 ($\pm 0.2$) | heart rate, injury time, respiratory, capillary refill time, systolic blood pressure |
| Shapley | 3.5 ($\pm 0.1$) | Injury type, gender, GCS, injury time, age |
| IG | 3.5 ($\pm 0.2$) | Injury type, gender, injury time, age |
| IST-3 |  |  |
| Saliency | 19.1 ($\pm 0.64$) | gcs, nihss, age, weight, infarct: possibly yes |
| SmoothGrad | 18.8 ($\pm 0.60$) | diastolic blood pressure, nihss, age, weight, gender |
| **Shapley** | **17.0 ($\pm 0.64$)** | **Nihss, infarct, anti-platelet usage, atrial fibrillation, gender** |
| IG | 17.0 ($\pm 0.66$) | Nihss, infarct, anti-platelet usage, atrial fibrillation, TACI |
| SPRINT |  |  |
| Saliency | 6.1 ($\pm 0.21$) | Umacr, EGFR, TRR, creatine, glucose |
| SmoothGrad | 5.9 ($\pm 0.21$) | creatinine, EGFR, age, diastolic blood pressure |
| Shapley | 5.1 ($\pm 0.12$) | CKD history, statin usage, Age, Cardiovascular history, gender |
| **IG** | **5.3 ($\pm 0.17$)** | **Age, gender, statin, CKD history, Cardiovascular history** |
| ACCORD |  |  |
| Saliency | 17.6 ($\pm 0.64$) | Trig, EGFR, Potassium, ALT, HDL |
| SmoothGrad | 17.0 ($\pm 0.64$) | HDL, potassium, EGFR, alt, Trig |
| **Shapley** | **14.7 ($\pm 0.56$)** | **Cardiovascular history, gender, aspirin, # of anti-bp agents, race** |
| IG | 14.8 ($\pm 0.57$) | Cardiovascular history, gender, aspirin, # of anti-bp agents, race |

### D.5 Global Sufficiency - Alignment between Reported Features and Shapley values

**Table S.10.**
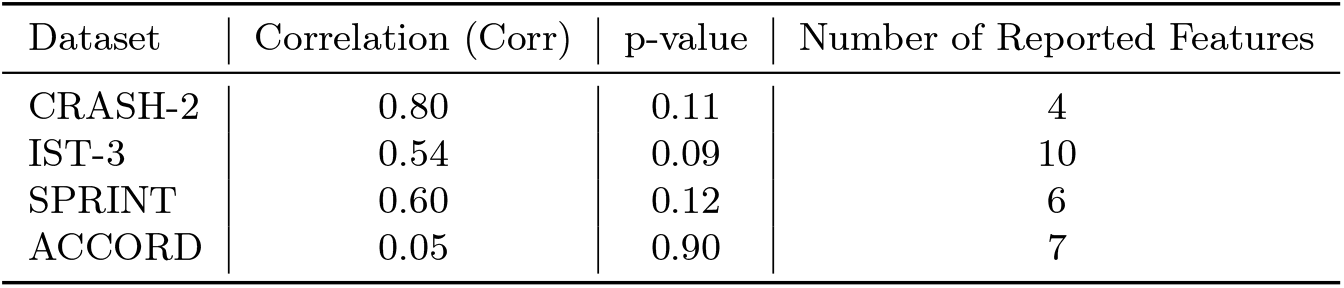
Correlation between ranks based on LIFT-XAI and interaction p-values across RCTs.

#### D.5.1 ACCORD & SPRINT Additional Results

**Table S.11.** Uplift and Qini scores with 95% confidence intervals for various datasets. (*) denotes datasets excluding individuals with glucose levels greater than 300 mg/dL in ACCORD and patients older than 45 y/o in the Harborview trauma registry.

| Dataset | Uplift Score ($\times 10^{-2}$) | Qini Score ($\times 10^{-2}$) |
| --- | --- | --- |
| SPRINT (train) | $7.5 \pm 0.11$ | $3.9 \pm 0.06$ |
| ACCORD (test) | $0.38 \pm 0.17$ | $0.22 \pm 0.09$ |
| ACCORD* | $0.53 \pm 0.17$ | $0.30 \pm 0.09$ |
| CRASH-2 (train) | $7.6 \pm 0.11$ | $4.0 \pm 0.07$ |
| Harborview (test) | $0.05 \pm 0.04$ | $-0.05 \pm 0.03$ |
| Harborview* | $0.50 \pm 0.08$ | $0.08 \pm 0.05$ |

### D.6 Pre-hospital (UW Harborview) & In-hospital (CRASH-2) TXA Additional Results

**Fig. S.5.**
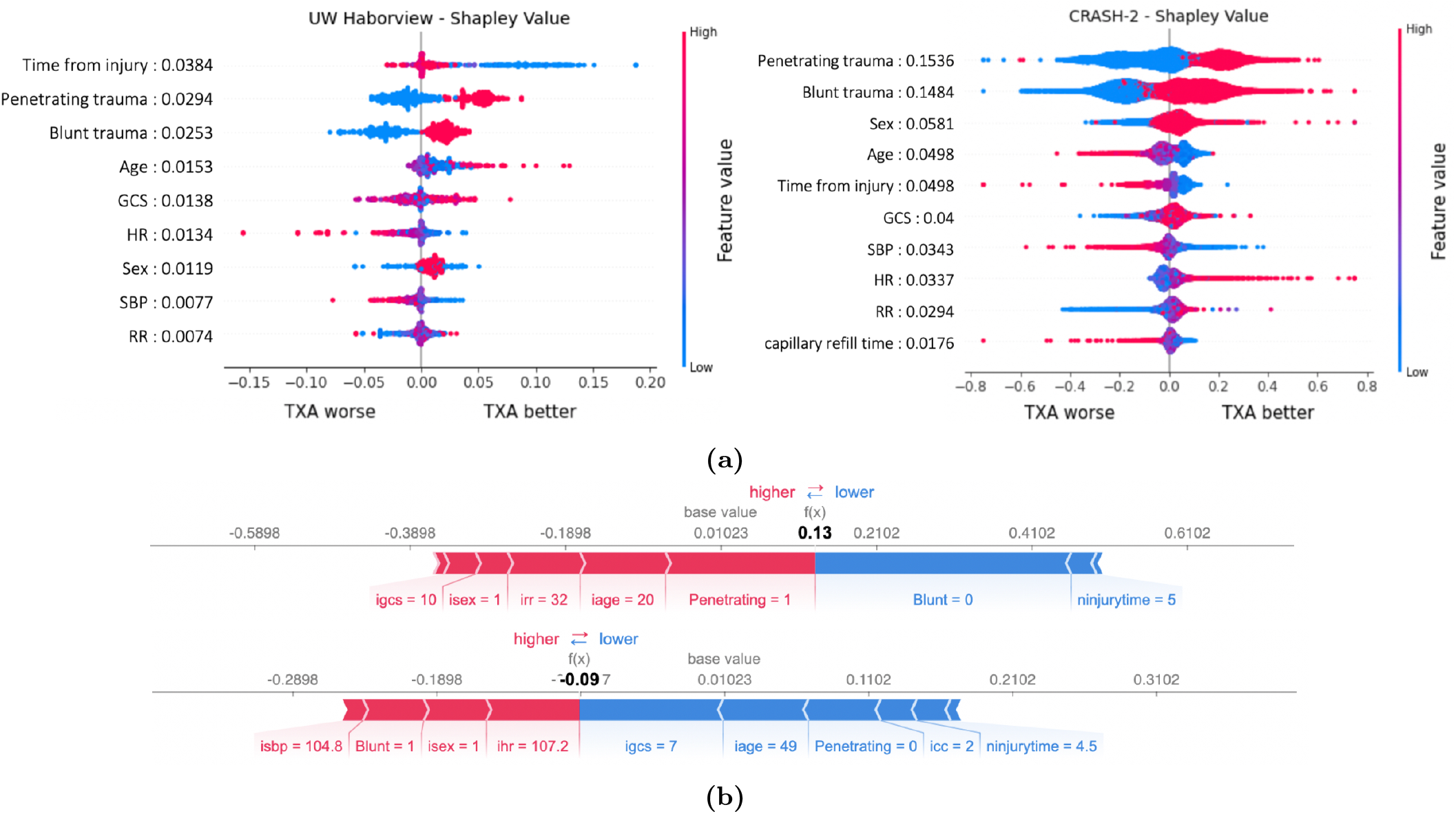
**(a)** Shapley summary plot for the Harborview cohort (left), CRASH-2 (right).**(b)** CRASH-2: Explaining sample individuals with different demographics and laboratory values. Red colors denote positive attributions and blue denotes negative attributions.

## E Agreement Between Physicians and LLM Judges in Logic-Gating Evaluation

### E.1 Specialist Characteristics

**Table S.12.** Distribution of clinical evaluators by specialty and attending experience across datasets.

| Dataset | Total | Attending (< 5 years) | Attending ($\geq 10$ years) |
| --- | --- | --- | --- |
| CRASH-2 (Emergency Medicine) | 2 | 0 | 2 |
| ACCORD (Cardiology) | 7 | 2 | 5 |
| ACCORD (Endocrinology & Metabolism) | 5 | 5 | 0 |
| IST-3 (Neurology) | 3 | 2 | 1 |
| SPRINT (Internal Medicine) | 2 | 2 | 0 |

### E.2 Inter-Rater Agreement

**Fig. S.6.**
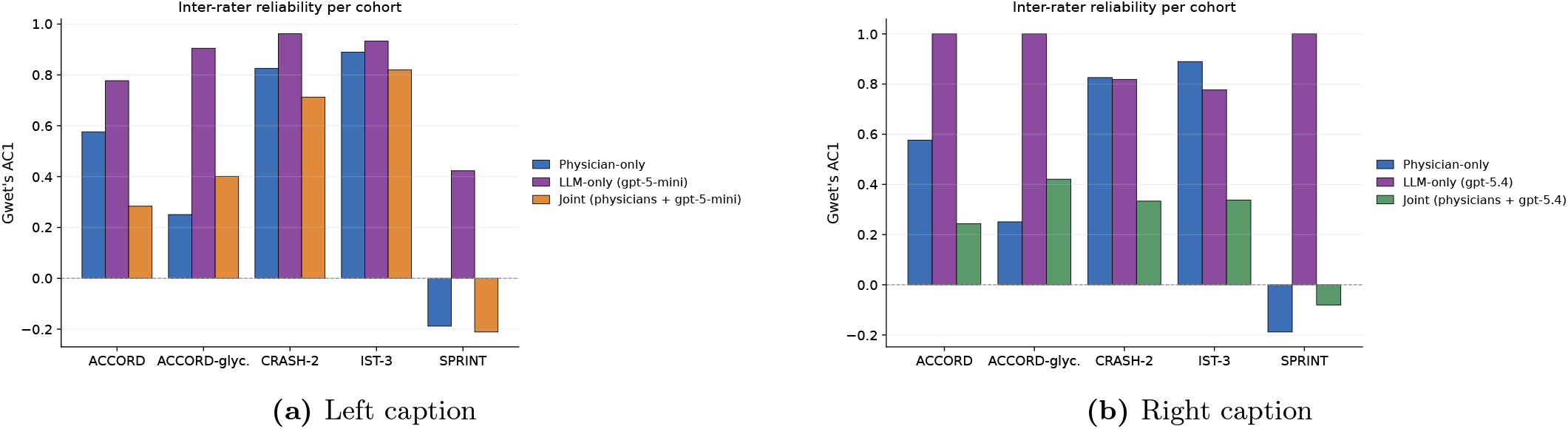
Inter-rater agreement between LLM judges and clinical specialists across cohorts, measured using Gwet’s AC1. Panels show results for (a) GPT-5-mini and (b) GPT-5.4.

## F Automated PubMed evidence retrieval

**Table S.13.**
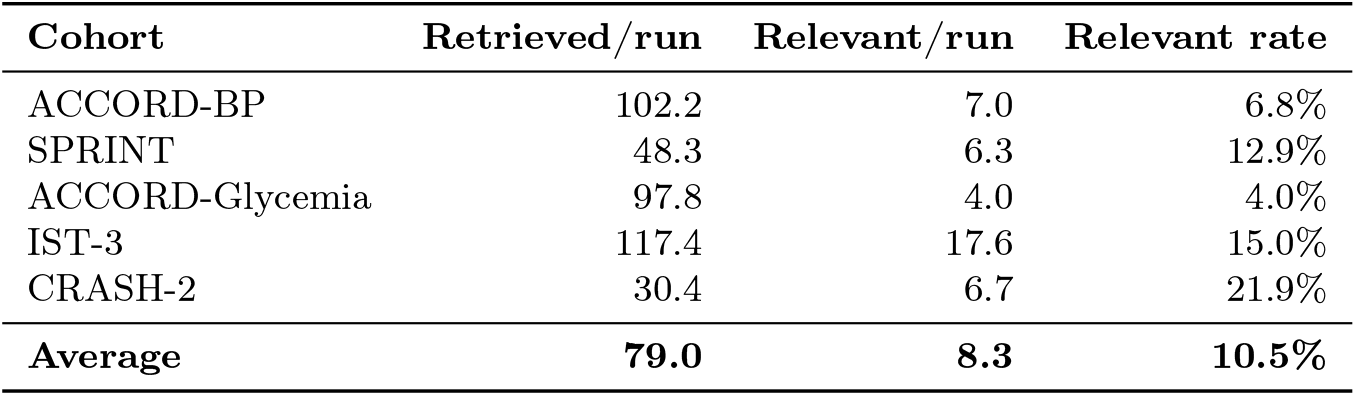
PubMed retrieval volume and relevance by dataset. Values are averaged across explanation-generation methods and random seeds. A run denotes one method–seed pair, with five runs performed per method. Relevant abstracts are those classified by the LLM judge as non-irrelevant after passing the Step-1 relevance gates.

**Table S.14.** Agreement between LLM-assigned evidence labels and human annotations for 125 sampled retrieved articles across cohorts. Precision, recall and F1 are computed using human annotations as the reference standard.

| Evidence label | Precision | Recall | F1 | Support |
| --- | --- | --- | --- | --- |
| Conflict | 1.00 | 1.00 | 1.00 | 5 |
| Irrelevant | 1.00 | 0.96 | 0.98 | 98 |
| No interaction | 0.62 | 0.83 | 0.71 | 6 |
| Prognostic main effect | 0.62 | 1.00 | 0.77 | 5 |
| Support interaction | 0.86 | 0.75 | 0.80 | 8 |
| Weak support | 1.00 | 1.00 | 1.00 | 3 |
| Exact agreement |  |  |  | 94.3% |
| Cohen's $\kappa$ | | | | 0.85 |

## G Example Output from ALEX

### G.1 Single Cohort Analysis ACCO

#### ACCORD-BP

1. **DBP** Patients with low baseline DBP derive less net benefit — or may be harmed — from an intensive SBP target because aggressive lowering further reduces diastolic pressure and compromises coronary perfusion (J-curve). The hypothesis is conditionally grounded: harm is most pronounced when low DBP co-occurs with established coronary artery disease, whereas patients with adequate DBP tolerate intensive SBP lowering with larger net benefit. Supported by J-curve evidence in observational data and INVEST subgroup analyses.
2. **SBP** Higher baseline SBP predicts greater absolute benefit because the achievable SBP reduction is larger and baseline risk of BP-sensitive events (stroke, HF) is higher. This is backed by the BPLTTC pooled analysis and explains differential results between ACCORD-BP (mixed baseline SBP, no overall benefit) and SPRINT (higher baseline SBP, clear benefit).

#### SPRINT

1. **CKD** Patients with mild-to-moderate CKD derive greater absolute CV-event reduction because of higher baseline event rates, but those with advanced CKD are at disproportionate risk of AKI and GFR decline that blunts net delivered benefit. This directly mirrors SPRINT’s own documented findings on renal adverse events and is well-grounded in hemodynamic autoregulation physiology.
2. **Age** Non-frail older adults (e.g., ≥ 75) benefit more in absolute terms due to higher baseline incidence of HF and stroke — consistent with SPRINT’s ≥ 75 subgroup result — while frail elders with competing mortality show attenuated net benefit. The hypothesis correctly conditions on frailty and comorbidity as moderators, which is the precise conceptual distinction that SPRINT investigators highlighted.

#### IST-3

1. **NIHSS** Moderate stroke severity (NIHSS ∼ 5–15) produces the largest absolute functional benefit because these patients have substantial salvageable penumbra without high baseline sICH risk; very mild NIHSS leaves minimal room for improvement, and very severe NIHSS carries high hemorrhagic risk and established infarct core that attenuates reperfusion benefit. This is the best-supported heterogeneity finding in thrombolysis meta-analyses (Emberson et al.).
2. **Glucose** Elevated admission glucose attenuates alteplase benefit by amplifying blood-brain barrier disruption, reperfusion injury, and hemorrhagic transformation — reducing functional gain per unit of recanalization. The interaction is conditional on whether glucose reflects chronic diabetes vs. acute stress hyperglycemia. Strong experimental and clinical plausibility.

#### CRASH-2

1. **Time-to-treatment** Treatment within the first 1–3 hours (especially the first hour) produces larger mortality reductions because early antifibrinolysis prevents progression of coagulopathy; beyond ∼ 3 hours, fibrinolytic activity has resolved or been replaced by secondary injury processes where TXA adds little or causes harm. This is the strongest and most directly validated finding from the original CRASH-2 time-stratified analysis.
2. **Injury type** Blunt polytrauma — which triggers systemic hyperfibrinolysis — confers greater TXA benefit than isolated focal penetrating injuries (where surgical hemostasis, not antifibrinolysis, dominates). Mechanistically, the benefit scales with the degree of trauma-induced coagulopathy, consistent with trauma physiology and CRASH-2’s population-level findings.

#### ACCORD Glycemia

1. **Baseline HbA1c** Patients with high baseline HbA1c derive less benefit or net harm from intensive glycemic targets because achieving HbA1c *<* 6.0% from a high starting point requires aggressive insulin/secretagogue escalation, producing more severe hypoglycemia that triggers ischemia and arrhythmias offsetting any atherosclerotic gains. The interaction is strongest when combined with long diabetes duration and rapid pharmacologic escalation — directly aligned with the mechanism suspected for excess mortality in ACCORD’s intensive arm.
2. **BP medication** Antihypertensive use (particularly beta-blockers) attenuates the benefit of intensive glycemic control by blunting hypoglycemia recognition and impairing compensatory adrenergic responses, increasing clinically significant hypoglycemic events. Patients on ≥ 2 BP agents also tend to have more advanced vascular disease (less reversible by glycemic lowering), so iatrogenic risk outweighs atherosclerotic benefit. This is a mechanistically coherent joint-drug interaction consistent with ACCORD’s pharmacology.

### G.2 Cross Cohort Analysis

#### Cross Cohort Analysis: ACCORD-SPRINT (BP)

1. **Fasting Plasma Glucose (FPG)** Patients with higher fasting plasma glucose derive greater absolute benefit from intensive SBP lowering because hyperglycemia amplifies pressure-mediated endothelial injury and microvascular dysfunction — meaning the same reduction in SBP produces a larger reduction in cerebrovascular and microvascular events in those with worse glycemic control. This is mechanistically intuitive: high glucose and high BP act synergistically to damage the endothelium, so lowering BP in that milieu has a greater impact. However, the interaction is conditional on renal function: high-FPG patients with preserved eGFR are most likely to realize this benefit, while those with advanced CKD or autonomic neuropathy face a greater risk of hypotension, AKI, and orthostatic adverse events that offset the gain. This conditionality is clinically important given that enrolled patients with T2DM, many of whom have concurrent CKD. Notably, ACCORD assigned patients to both a BP arm and a glycemia arm — so the observed FPG × BP-treatment interaction needs to be estimated adjusting for glycemic arm assignment to avoid confounding between the two factorial interventions.

#### Cross Cohort Analysis: CRASH-2-Harborview (TXA)

1. **Age** Older patients derive less absolute survival benefit from pre-hospital TXA due to age-related shifts in baseline hemostasis. Chronological aging is associated with baseline endothelial dysfunction and elevated endogenous antifibrinolytics (e.g., PAI-1), resulting in a blunted hyperfibrinolytic response to trauma-induced shock. Consequently, older patients have a smaller mechanistic target for exogenous TXA compared to younger patients; furthermore, TXA may disproportionately tip the older patient’s already pro-thrombotic baseline state into microvascular thrombosis, attenuating net survival benefit. Caveat: The interaction is conditional on the actual coagulation phenotype at presentation. Older patients who do develop severe hyperfibrinolysis will still benefit, but at a population level, the age-related shift in baseline coagulation profiles attenuates the average absolute benefit of empiric TXA administration.

## H Prompt Template

### H.1 Prompt Template for ALEX Prompt

#### Prompt for XAI Agent

You are an XAI (Explainable AI) research agent for clinical trial data. You have access to tools that let you:

1. Inspect a dataset (`load_dataset`)
2. Train a CATE model and compute ensemble Shapley values (`run_shap_pipeline`)
3. Load an existing SHAP summary without recomputing (`load_shap_summary`)
4. Generate mechanistic hypotheses explaining the SHAP results (`generate_hypotheses`)

Work step-by-step. Inspect the data first, then compute or load SHAP values, then optionally generate hypotheses. When you are done, provide a concise written summary of your findings: which features drive treatment heterogeneity and what that implies clinically.

#### Prompt for Generation Agent

Prompt for Generation Agent

Act as a Clinical Expert analyzing treatment effect heterogeneity. You must exclusively focus on predictive factors (effect modifiers)—features that change the magnitude or direction of the treatment’s benefit. You must completely ignore purely prognostic factors—features that merely predict the baseline outcome regardless of treatment—unless they actively alter the drug’s efficacy.

You are analyzing feature attribution from a conditional average treatment effect model using trial metadata (treatment/outcome/population).

Generate explanations on why these specific features could modify treatment response. Treat SHAP-ranked features as model signals to interpret. If a feature is coded/ambiguous or not strongly established in literature, keep it and explain it as a plausible proxy or exploratory/novel modifier with explicit uncertainty. For each feature, propose distinct hypotheses/explanations explaining how it modifies the treatment effect. Each explanation should offer a different perspective or mechanism (e.g., direct biological effect, proxy/confounding role, pharmacological interaction).

Please follow the following requirements:

1. Focus STRICTLY on Heterogeneous Treatment Effects (Interaction), not just main effects.
2. If a feature has a bidirectional effect (e.g., good for some, bad for others), specify that.
3. Grounding: Please follow known physiological/biological principles.
4. For each feature, distinguish whether support is established vs exploratory; include caveats when evidence is limited rather than dropping the feature.
5. Every mechanism description MUST be framed as a treatment-response interaction, NOT a prognostic statement. For example, ‘Older patients show attenuated benefit from chemotherapy due to reduced tolerability and higher rates of dose-limiting adverse events, resulting in smaller net absolute benefit compared to younger patients with equivalent disease burden.’ Always specify: who benefits more/less, from which treatment, and why the magnitude of benefit differs between subgroups.
6. Avoid oversimplified monotone subgroup claims: Real treatment-effect heterogeneity is often conditional on multiple factors. If a feature’s interaction with treatment depends on a second moderator (e.g., age × comorbidity, eGFR × diabetes duration), name that conditionality explicitly rather than stating a blanket directional claim. For example, ‘Among patients aged ≥ 75 WITHOUT established CVD, statin therapy yields smaller absolute benefit due to competing mortality risks and shorter life expectancy; however, among older patients WITH pre-existing CVD, statin treatment still substantially reduces cardiovascular events.’

#### Prompt for Verifier Agent

Prompt for Verifier Agent

You are a collaborative scientific advisor helping to refine feature-level mechanistic explanation. Your role is to improve the explanations, not just evaluate them. For each explanation: 1) Identify strengths worth preserving, 2) Spot weaknesses that need improvement, 3) Provide constructive refinement suggestions, and 4) ALWAYS return a revised, improved version. Focus on

- Strengthening mechanism plausibility with more specific biological/clinical details
- Sharpening clinical interpretation to be more precise
- Better aligning with SHAP evidence (direction, magnitude)
- Making subgroup implications more actionable
- Enhancing validation plans with concrete, feasible steps
- Adding important caveats and alternative explanations
- Rewrite any mechanism description that is framed as a prognostic statement into a treatment-response interaction claim. Every description must answer: which subgroup benefits more (or less) from treatment, and why the magnitude of treatment effect differs. For example, ‘Patients with high baseline LDL (*>* 190 mg/dL) derive greater absolute benefit from statin therapy because their elevated baseline event rate translates even a moderate relative risk reduction into a larger absolute risk reduction per patient treated.’ If a description does not specify differential treatment response, rewrite it so it does.
- Flag and rewrite any oversimplified monotone subgroup claim. If the feature’s interaction with treatment is moderated by a second factor (e.g., age comorbidity, eGFR diabetes duration, LDL × CVD history), the refined description must capture that conditionality explicitly rather than stating a blanket direction.

For EACH explanation:

- Assess plausibility (high/moderate/low/implausible)
- Suggest specific improvements to the description
- Mark for revision if implausible or poorly supported

If trial article context provided:

- Refine mechanisms to align with known trial physiology
- Adjust interpretations to match population characteristics
- Tailor validation plans to be feasible within trial design
- Flag any contradictions with trial findings

Your goal is to help create the BEST possible explanations. Always provide a complete revised FeatureHypothesesSet with improvements, even if changes are minor. Build on strengths and fix weaknesses. Stay grounded in evidence - improve but don’t add unsupported claims. You MUST preserve ALL explanations for every feature. Never reduce the number of hypotheses. If the input has 2 hypotheses for a feature, the output must also have exactly 2 hypotheses. Refine or rewrite descriptions, but do NOT drop or omit any.

### H.2 Prompt Template for Judges

#### Prompt for Judge Agent in Logic Gating Evaluation

You are an independent scientific judge for mechanistic hypotheses. Evaluate each explanation using a strict cascading gate framework. Be objective, fair, and constructive.

Score each explanation entry *individually*. Multiple entries may share the same feature name but correspond to different subgroup explanations. Copy the `explanation_id` for each input entry so every score can be traced back to its source explanation.

**Evaluation process (follow in order for each explanation):**

1. Write `gate_reasoning`: one sentence per gate (G1–G5) before assigning booleans.
  - G2: ask whether a concrete biological pathway is stated (for example, receptor, enzyme, PK/PD change), not merely a statement that higher baseline risk implies larger benefit.
  - G3: ask whether the drug truly acts differently in this subgroup, beyond pure absolute-risk amplification. Confirm that the feature is baseline, not post-treatment, and not reverse-causal.
  - If a gate fails, explicitly state: `G[N] FALSE –-cascade: G[N+1..5] also FALSE`.
2. Set each boolean consistently with the gate reasoning.
3. Write a one-sentence `justification` naming the decisive gate and why.

**Cascading robustness gates:** Each gate is binary and must be evaluated in order. A explanation that fails an earlier gate cannot receive credit for later gates. The overall score is:

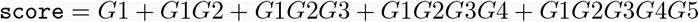

**Gate 1 — Observed in data (**`is_observed_in_data`)

True if the proposed modifying feature, or a close synonym, appears in the authoritative list of trial-measured features. False if the feature is absent, inferred post hoc, or derived from unavailable data. If false, all later gates must also be false.

**Gate 2 — Is logically coherent (**`is_logically_coherent`)

True only if the explanation describes a concrete, coherent rationale. For example, a biological mechanism connecting the feature to differential drug action, such as a receptor, enzyme, pharmacokinetic/pharmacodynamic shift, or cell pathway. False if the argument is purely statistical, circular, tautological, vague, or biologically implausible.

**Gate 3 — Causally plausible (**`is_causally_plausible`)

True only if the feature is a genuine treatment-effect modifier, meaning the drug works differently in this subgroup because of a specific mechanistic pathway. False if the claim reflects only absolute-risk amplification, a post-randomization or post-treatment variable, reverse causality, or a trivial severity proxy without additive mechanism.

**Gate 4 — Clinically actionable (**`is_clinically_actionable`)

True if the explanation defines clear and operationalizable patient subgroups with distinct treatment implications for real-world care. False if the subgroup is vague, impractical, or clinically insignificant.

**Gate 5 — Literature backed (**`is_literature_backed`)

True if the specific feature-by-treatment interaction is supported by published clinical literature, ideally sub-group analyses, randomized trials, or meta-analyses. False if speculative, contradicted, or unsupported.

**Novelty bonus (**`is_novel`)

Mark true if the explanation identifies an underexplored mechanism or subgroup not already well covered in major reviews or guidelines. This does not affect the overall score.

**Use of trial context:** If trial article context is provided, use it to verify whether features were measured, whether the proposed pathway is biologically coherent, whether the feature is a genuine modifier rather than a proxy, and whether subgroup interactions are supported by the original trial report. Provide honest, rigorous judgments. Default to false when uncertain.

### H.3 Prompt Template for Article Retrieve Prompt for PubMed Article Retrieval

#### Prompt for PubMed Article Retrieval

You are a clinical librarian building a PubMed search strategy.

1. **Trial Context (PICO frame)**
  - **P — Population:** [POPULATION]
  - **I — Intervention:** [TREATMENT]
  - **C — Comparator:** [COMPARATOR]
  - **O — Outcome:** [OUTCOME]
2. **Effect Modifier Under Test**
  - **Feature:** “[TARGET FEATURE]”
  - **Hypothesised mechanism:** [MECHANISM DESCRIPTION]
  - **Expected direction:** [EXPECTED DIRECTION]
  - **Known clinical aliases** (when the variable name is opaque, e.g. `icc`, `sub_cvd`): [ALIAS LIST] These describe what the variable *actually* measures clinically; the raw variable name may be a code (e.g. “icc” in trauma cohorts means capillary refill, NOT ICD code; “sub_cvd” means history of CVD, NOT a CVD subgroup analysis). `feature_concepts` MUST include these aliases.
3. **Task** Produce a search strategy that retrieves RCTs, secondary analyses, and meta-analyses likely to report whether “[TARGET FEATURE]” modifies the treatment effect of **I** versus **C** on **O** in **P**. Do **not** guess MeSH descriptor spellings. The system canonicalises your concepts to MeSH automatically via PubMed’s TranslationSet. Emit clinical concepts in plain English.
4. **Output — STRICT JSON with the following keys**
  - `treatment_concepts` — **I**: drug or intervention names and clinical synonyms.
  - `population_concepts` — **P**: disease/population names, e.g. “ischemic stroke”.
  - `feature_concepts` — the effect-modifier concept and its synonyms.
  - `interaction_terms` — phrasing that signals subgroup or HTE analysis, e.g. “effect modification”, “interaction p”, “predictive factor”.
  - `required_keywords` — ≥ 1 must appear in title+abstract; cheap pre-screen.
  - `excluded_keywords` — disqualifying tokens, e.g. wrong disease, population, or drug.
  - `competing_interventions` — drugs/devices that act as the *primary* arm in studies where **I** is only the comparator.
  - `expected_yield` — integer in [5, 200].
5. **Rules**
  - Concepts must be standard clinical English; the system maps to MeSH.
  - `excluded_keywords` must not overlap any `*_concepts` or `required_keywords`.
  - `competing_interventions` must not overlap `treatment_concepts`.
  - Keep each list ≤ 12 items. Prefer specific over generic.
  - Output JSON only — no commentary.
6. **Downstream pipeline (deterministic, no LLM)**
  - **MeSH canonicalisation**. Each concept is mapped to MeSH descriptors via PubMed’s TranslationSet, filtered by token overlap with the source concept, and capped at the top 2 descriptors per concept.
  - **Tiered queries**. Up to five PubMed queries are compiled in parallel: LLM-strict with interaction terms, LLM-no-interaction, and three curated tiers from v1: mechanism, strict, and broad. Each is wrapped with publication-type filters: `Clinical Trial`, `Randomized Controlled Trial`, `Meta-Analysis`, and `Review`.
  - **Competing-intervention exclusion**. A title-only `NOT` clause excludes papers whose *title* centres on a competing intervention.
  - **Reciprocal-Rank Fusion (RRF)**. Per-tier ranked PMID lists are fused with score(*p*) = ∑_*t*_ 1*/*(*k* + *r*_*t*_(*p*) + 1), where *k* = 20; the top-*K* PMIDs proceed.
  - **Title-only prescreen**. Abstracts whose title contains any `excluded_keyword` are dropped before the judge using word-boundary regex, after stripping evidence-type tokens such as “retrospective” or “nonrandomized”.

### H.4 Prompt Template for PubMed Validator Prompt for PubMed Validator

#### Prompt for PubMed Validator

You are an expert biostatistician and medical researcher. Your task is to evaluate if the Clinical Abstract provides evidence supporting the proposed explanation/hypothesis/mechanism.

1. The explanation/hypothesis/mechanism:
  - **Trial Context:** [DATASET] ([TREATMENT] vs [COMPARATOR])
  - **Outcome:** [OUTCOME]
  - **TARGET FEATURE:** [TARGET FEATURE]
  - **Mechanism:** [MECHANISM DESCRIPTION]
  - **Expected Direction:** [EXPECTED DIRECTION]
2. The abstract:
  - **Title:** [ABSTRACT TITLE]
  - **Text:** [ABSTRACT TEXT]
3. **STEP 1: RELEVANCE GATES (Pass/Fail)**
  - **(A) Treatment Check:** Is [TREATMENT] (or class equivalent) evaluated?
  - **(B) Outcome Alignment:** Must evaluate ‘[OUTCOME]’ or surrogate.
  - **(C) Feature Check:** Is [TARGET FEATURE] explicitly analyzed for outcome association?
    - – REJECT if feature is missing or only a covariate/baseline stat not linked to outcome.
  - **(D) Intervention Match:** The abstract must test [TREATMENT] as a PRIMARY arm.
    - – REJECT if the study primarily evaluates a DIFFERENT intervention that happens to use [TREATMENT] as a control.
  - **(E) Population Context:** The study population must be reasonably comparable to [POPULATION].
    - – REJECT if the abstract studies a fundamentally DIFFERENT disease entity, even if the same drug is used.
    - *– Example REJECT:* A study evaluating the treatment in a fundamentally different pathophysiological condition is IRRELEVANT.
    - *– Example PASS:* A study in a related condition may be relevant if the physiological dynamics are analogous and the authors explicitly discuss generalizability.
    - → If (A), (B), (C), (D), or (E) fails, output: **[F] IRRELEVANT**.
4. **STEP 2: EVIDENCE EVALUATION (The “Mechanism Test”)** If the abstract passes Step 1, compare the REPORTED RESULTS against the EXPECTED CLINICAL OUTCOME.
  - **[A] SUPPORT INTERACTION (Mechanism Supported):** Explicit statement that the magnitude of treatment benefit is SIGNIFICANTLY DIFFERENT between subgroups defined by [TARGET FEATURE]. Directionality must match. Includes numeric evidence (e.g., Interaction *p <* 0.05) or strong textual claims. *ABSOLUTE vs RELATIVE SCALE:* Evidence of greater absolute risk reduction counts even if relative risk ratio is consistent.
  - **[B] SUPPORT WEAK (Mechanism Consistent):** Numerical trend or subgroup observation favoring the mechanism, but explicitly noted as not statistically significant (e.g., *p >* 0.05) or hypothesis-generating.
  - **[C] PROGNOSTIC MAIN EFFECT (Mechanism Unclear):** Evidence that the feature predicts the Outcome generally, but NOT the response to the specific drug.
  - **[D] NO INTERACTION (Mechanism Inactive):** Explicit statement that the feature does NOT modify the treatment effect (e.g., “Outcomes were similar regardless of status”).
  - **[E] CONFLICT (Mechanism Contradicted):** Significant Interaction in the OPPOSITE direction of the hypothesis.
5. **STEP 3: STUDY DESIGN CLASSIFICATION** Classify the study based solely on the abstract: `RCT`, `RCT_secondary`, `systematic_review_meta_analysis`, `prospective_cohort`, `retrospective_cohort`, `case_control`, `cross_sectional`, `case_series`, `narrative_review`, or `other`.
6. **OUTPUT FORMAT:** Return a valid JSON object with the following fields: `classification`, `confidence`, `study_design`, `reasoning`, and `evidence_quote`.

## Footnotes

1 https://github.com/AliciaCurth/CATENets

2 https://captum.ai/

3 https://github.com/AliciaCurth/CATENets

4 https://github.com/jpaillard/permucate/blob/main/scripts/ihdp_exp.py

5 Note that this is different from the random treatment assignment in RCT.

6 https://captum.ai/

7 For local attribution methods, global rankings are obtained by averaging absolute attributions across individuals.

8 This procedure is conceptually similar to RemOve And Retrain (ROAR) ^56^, but evaluates explanations at the global level.

